# REGEN-COV Antibody Cocktail Clinical Outcomes Study in Covid-19 Outpatients

**DOI:** 10.1101/2021.05.19.21257469

**Authors:** David M. Weinreich, Sumathi Sivapalasingam, Thomas Norton, Shazia Ali, Haitao Gao, Rafia Bhore, Jing Xiao, Andrea T. Hooper, Jennifer D. Hamilton, Bret J. Musser, Diana Rofail, Mohamed Hussein, Joseph Im, Dominique Y. Atmodjo, Christina Perry, Cynthia Pan, Adnan Mahmood, Romana Hosain, John D. Davis, Kenneth C. Turner, Alina Baum, Christos A. Kyratsous, Yunji Kim, Amanda Cook, Wendy Kampman, Lilia Roque-Guerrero, Gerard Acloque, Hessam Aazami, Kevin Cannon, J. Abraham Simón-Campos, Joseph A. Bocchini, Bari Kowal, Thomas DiCioccio, Yuhwen Soo, Neil Stahl, Leah Lipsich, Ned Braunstein, Gary Herman, George D. Yancopoulos, for the Trial Investigators

## Abstract

**Background:** REGEN-COV antibody cocktail (casirivimab with imdevimab) rapidly reduced viral load and decreased medically-attended visits in the phase 1/2 portion of this trial; REGEN-COV, retains activity *in vitro* against emerging SARS-CoV-2 variants of concern.

**Methods:** The phase 3 portion of this adaptive, randomized, master protocol, included 4,057 Covid-19 outpatients with one or more risk factors for severe disease. Patients were randomized to a single treatment of intravenous placebo, or various doses of REGEN-COV, and followed for 28 days. The prespecified hierarchical analysis first compared REGEN-COV 2400mg dose vs concurrent placebo, then compared the 1200mg dose vs concurrent placebo, for endpoints assessing risk of hospitalization or death, and time to symptom resolution. Safety was evaluated in all treated patients.

**Results:** Both REGEN-COV 2400mg and 1200mg significantly reduced Covid-19-related hospitalization or all-cause death compared to placebo (71.3% reduction [1.3% vs 4.6%; p<0.0001] and 70.4% reduction [1.0% vs 3.2%; p=0.0024], respectively). The median time to resolution of Covid-19 symptoms was 4 days shorter in both dose arms vs placebo (10 vs 14 days; p<0.0001). Efficacy of REGEN-COV was consistent across subgroups, including patients who were SARS-CoV-2 serum antibody-positive at baseline. REGEN-COV more rapidly reduced viral load than placebo. Serious adverse events occurred more frequently in the placebo group (4.0%) than in the 1200mg (1.1%) and 2400mg (1.3%) groups and grade ≥2 infusion-related reactions were infrequent (<0.3% in all groups).

**Conclusions:** Treatment with REGEN-COV was well-tolerated and significantly reduced Covid-19-related hospitalization or all-cause death, rapidly resolved symptoms, and reduced viral load.

(Funded by Regeneron Pharmaceuticals and the Biomedical and Advanced Research and Development Authority of the Department of Health and Human Services; ClinicalTrials.gov number, NCT04425629.)

## INTRODUCTION

SARS-CoV-2 causes coronavirus disease 2019 (Covid-19) and, as of April 2021, has infected >130 million people and led to ∼3 million deaths globally.^1^ Although most Covid-19 patients are managed in the outpatient setting, some will progress to severe illness leading to hospitalization or death.^2-6^ Several investigational products, including REGEN-COV™, are available under Emergency Use Authorization but there have been limited clinical data to support their wider use and there are no approved treatments for reducing the risk of hospitalization or death due to Covid-19. There is also a need for therapeutics that remain effective against emerging SARS-CoV-2 variants of concern (VOCs), which contain mutations that attenuate immunity from prior SARS-CoV-2 infection, vaccination, and some monoclonal antibodies.^7-10^

To develop a therapeutic that would retain activity against emerging variants, a high-throughput screen was undertaken to generate an antibody cocktail consisting of two SARS-CoV-2 neutralizing antibodies against distinct, non-overlapping epitopes on the spike protein, resulting in REGEN-COV.^11-13^ In vitro studies demonstrated that REGEN-COV retains activity against current VOCs and variants of interest (VOIs), including B.1.1.7, B.1.429, and E484K-containing variants, such as B.1.351, P.1, and B.1.526.^9,14^ In the phase 1/2 portion of this adaptive phase 1-3 randomized, placebo-controlled master protocol, REGEN-COV demonstrated efficacy in symptomatic outpatients, where it rapidly reduced viral load and the need for medical attention related to Covid-19.^15,16^ On February 19^th^, 2021, an independent data monitoring committee (IDMC) recommended stopping enrollment of patients into the placebo group of the phase 3 portion of this master protocol because of clear efficacy of REGEN-COV. Here, we report the primary analysis results of the phase 3 portion of this trial.

## METHODS

### TRIAL DESIGN

This was an adaptive, multicenter, randomized, double-blind, placebo-controlled, phase 1/2/3 master protocol in Covid-19 outpatients (NCT04425629). The phase 3 portion comprised 3 cohorts: Cohort 1 (≥18 years), Cohort 2 (<18 years), and Cohort 3 (pregnant at randomization). Initially, phase 3 patients were randomized 1:1:1 to receive intravenous (IV) placebo, REGEN-COV 2400mg (1200mg each of casirivimab and imdevimab), or REGEN-COV 8000mg (4000mg each antibody) (Figure S1). Based upon phase 1/2 results that showed the 2400mg and 8000mg doses had similar antiviral and clinical efficacy and that most clinical events occurred in high-risk patients,^15^ the trial was amended on November 14^th^, 2020 so that subsequent patients enrolled had ≥1 risk factor for severe Covid-19 and were randomized 1:1:1 to receive placebo, REGEN-COV 1200mg (600mg each antibody) IV, or REGEN-COV 2400mg (1200mg each antibody) IV. On February 19^th^, 2021, per IDMC recommendation, patients were no longer randomized to receive placebo. The phase 3 primary efficacy analysis presented here is comprised of Cohort 1 patients randomized to REGEN-COV 2400mg or 1200mg, with their concurrent placebo groups serving as a control.

### TRIAL OVERSIGHT

Regeneron designed the trial; gathered the data, together with the trial investigators; and analyzed the data. Regeneron and the authors vouch for the accuracy and completeness of the data, and Regeneron vouches for the fidelity of the trial to the protocol. The authors provided critical feedback and final approval of the manuscript for submission.

The investigators, site personnel, and Regeneron employees who were involved in collecting and analyzing data were unaware of the treatment-group assignments. An independent data monitoring committee monitored unblinded data to make recommendations about trial modification and termination.

The trial was conducted in accordance with the principles of the Declaration of Helsinki, International Council for Harmonisation Good Clinical Practice guidelines, and applicable regulatory requirements. The local institutional review board or ethics committee at each study center oversaw trial conduct and documentation. All patients provided written informed consent before participating in the trial.

### PATIENTS

Eligible patients (Cohort 1) were ≥18 years of age and non-hospitalized, with a confirmed local SARS-CoV-2-positive diagnostic test result ≤72 hours and onset of any Covid-19 symptom, as determined by the investigator, ≤7 days before randomization. Randomization into the initial phase 3 portion was stratified by country and presence of risk factors for severe Covid-19. In the amended phase 3 portion, only patients with ≥1 risk factor for severe Covid-19 were eligible. The full list of inclusion and exclusion criteria can be found in the Protocol.

All patients were assessed at baseline for anti-SARS-CoV-2 antibodies – anti-spike [S1] IgA, anti-spike [S1] IgG, and anti-nucleocapsid IgG – and grouped for analyses as serum antibody–negative (if all available test results were negative), serum antibody– positive (if any available test result was positive), or other (inconclusive/unknown results).

### INTERVENTION AND ASSESSMENTS

At baseline (day 1), REGEN-COV (diluted in normal saline solution) or saline placebo was administered intravenously. Hospitalizations were assessed to be related to Covid-19 by the investigator. The Symptoms Evolution of COVID-19 (SE-C19) instrument, an electronic diary, recorded 23 Covid-19 symptoms daily.^17^ Quantitative virologic analysis of nasopharyngeal (NP) swab samples and serum antibody testing were conducted in a central laboratory and were previously described.^16^ Additional details on assessments are described in the Supplementary Appendix.

### ENDPOINTS

The primary and two key secondary endpoints were tested hierarchically (Table S1). The primary endpoint was the proportion of patients with ≥1 Covid-19-related hospitalization or all-cause death through day 29. The key secondary clinical endpoints were (1) the proportion of patients with ≥1 Covid-19-related hospitalization or all-cause death from day 4 through day 29 and (2) the time to Covid-19 symptoms resolution (19 of the 23 recorded symptoms). Time to Covid-19 symptoms resolution was defined as time from randomization to the first day during which the patient scored “no symptom’ (score=0) on all 19 symptoms except cough, fatigue, and headache, which could have been scored as “mild/moderate symptom” (score=1) or “no symptom” (score=0).

Safety endpoints for the phase 3 portion of the trial included serious adverse events (SAEs) that occurred or worsened during the observation period and adverse events of special interest (AESIs): grade ≥2 hypersensitivity reactions, grade ≥2 infusion-related reactions, and treatment-emergent adverse events (TEAEs) requiring medical attention at a healthcare facility.

### STATISTICAL ANALYSIS

The statistical analysis plan for the presented analysis was finalized prior to database lock and unblinding of phase 3 Cohort 1; the primary analysis did not include patients from the previously reported phase 1/2 portion of the trial.^16^ The full analysis set (FAS) included all randomized symptomatic patients. Efficacy analyses were performed based on a modified FAS (mFAS) defined as all randomized patients with a positive SARS-CoV-2 central lab-determined RT-qPCR test at baseline and with ≥1 risk factor for severe Covid-19. Safety was assessed in treated patients in the FAS.

The proportion of patients with ≥1 Covid-19-related hospitalization or all-cause death was compared between each dose group and concurrent placebo using the stratified Cochran-Mantel-Haenszel (CMH) test with country as a stratification factor. P-values from the stratified CMH test and 95% confidence intervals (CIs) for the relative risk reduction using the Farrington-Manning method are presented. Time to Covid-19 symptoms resolution was assessed in patients with a baseline total severity score >3 and analyzed using the stratified log-rank test with country as a stratification factor. Median times and associated 95% CIs were derived from the Kaplan-Meier method. The hazard ratio and 95% CI were estimated by the Cox regression model.

Analyses of the primary and key secondary endpoints were conducted at a two-sided α=0.05 utilizing a hierarchical testing strategy to control for type I error (Table S1). Statistical analyses were performed with SAS software, v9.4 (SAS Institute). Additional statistical methods are described in the Supplementary Appendix.

## RESULTS

### TRIAL POPULATION

Phase 3 patients were enrolled between September 24^th^, 2020 and January 17^th^, 2021. Initially, in the original phase 3 portion, a total of 3088 patients, with or without risk factors for severe Covid-19, were randomized to receive a single dose of either placebo, REGEN-COV 8000mg or REGEN-COV 2400mg. In the amended phase 3 portion, an additional 2519 patients with ≥1 risk factor were randomized to receive a single dose of placebo, REGEN-COV 2400mg or REGEN-COV 1200mg (Figure 1). Patients had a median follow-up of 45 days, with 96.6% of patients having >28 days follow-up.

**Figure 1.**
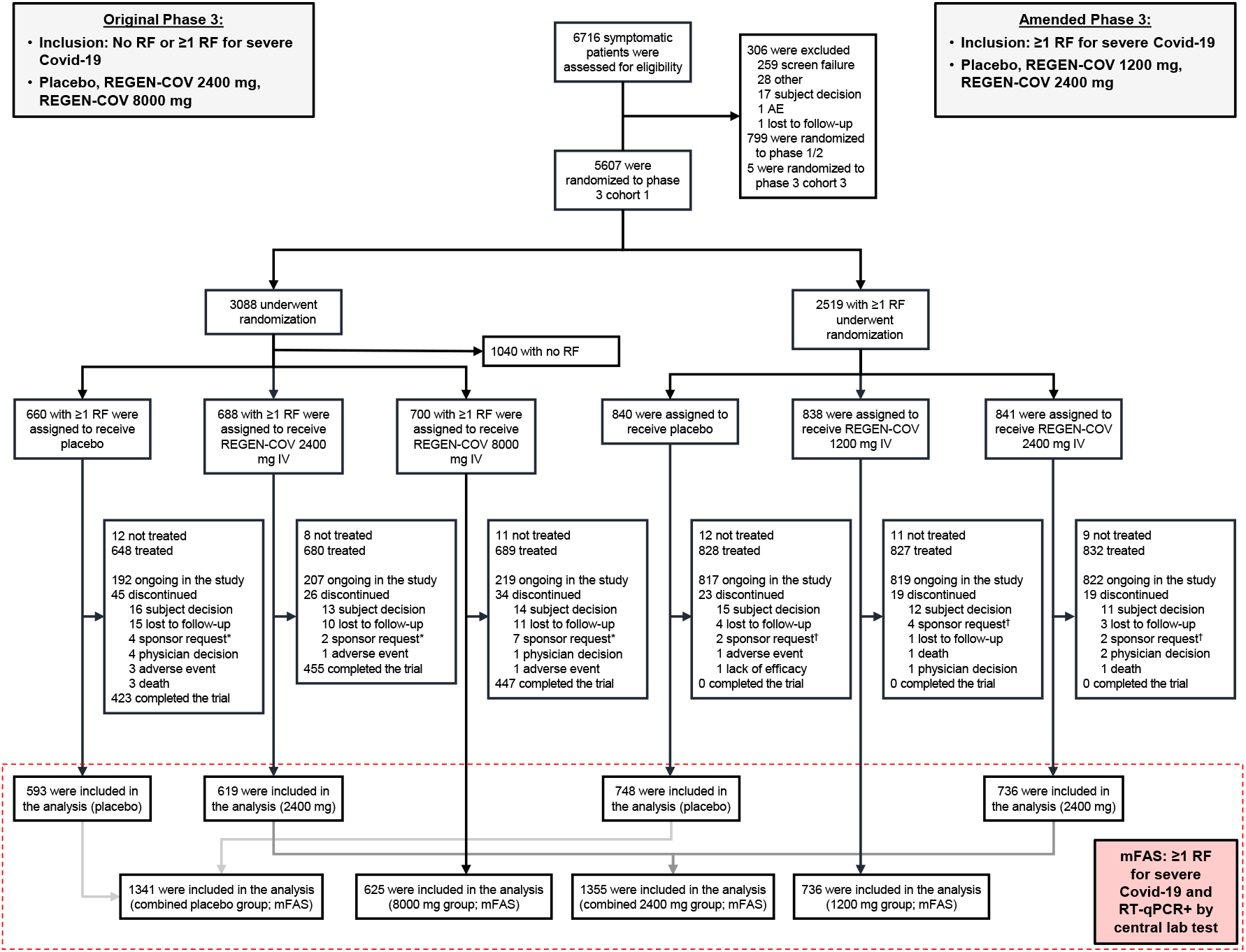
Screening, Randomization, Treatment, and Analysis. * In the original phase 3 portion of the trial, Regeneron requested that 2, 1, and 5 patients in the placebo, 2400 mg, and 8000 mg groups, respectively, withdraw from the trial as these patients did not meet the inclusion/exclusion criteria (ie, were randomized in error). Regeneron requested additional patients to withdraw from the trial due to site closure and inability to receive treatment. † In the amended phase 3 portion of the trial, Regeneron requested that 2, 4, and 2 patients in the placebo, 1200 mg, and 2400 mg groups, respectively, withdraw from the trial as these patients did not meet the inclusion/exclusion criteria (ie, were randomized in error). IV, intravenous(ly); mFAS, modified full analysis set; RF, risk factor; RT-qPCR, reverse transcriptase quantitative polymerase chain reaction.

The primary efficacy population included those with ≥1 risk factor for severe Covid-19 and a baseline central laboratory test positive for SARS-CoV-2 (mFAS) (Figure 1). Among the mFAS population (n=4057), demographic and baseline medical characteristics were balanced between the placebo and REGEN-COV groups (Table 1). For the overall mFAS population, median age was 50 years (interquartile range [IQR] 38-59), 49% were male, 14% ≥65 years, and 35% were Hispanic; the most common risk factors were obesity (58%), age ≥50 years (52%), and cardiovascular disease (36%); 3% of patients were immunosuppressed or on immunosuppressive medications (Table S2).

**Table 1.**
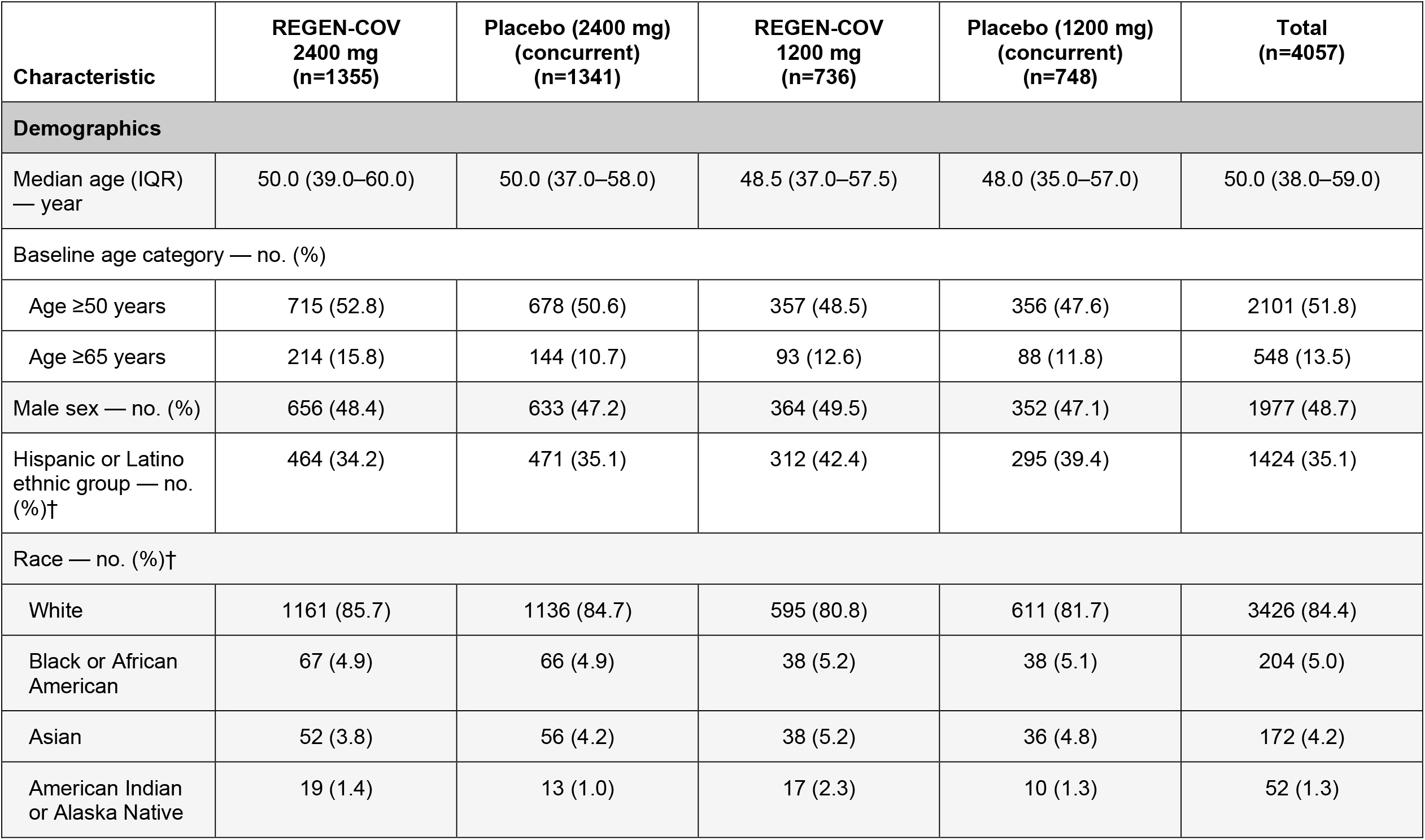
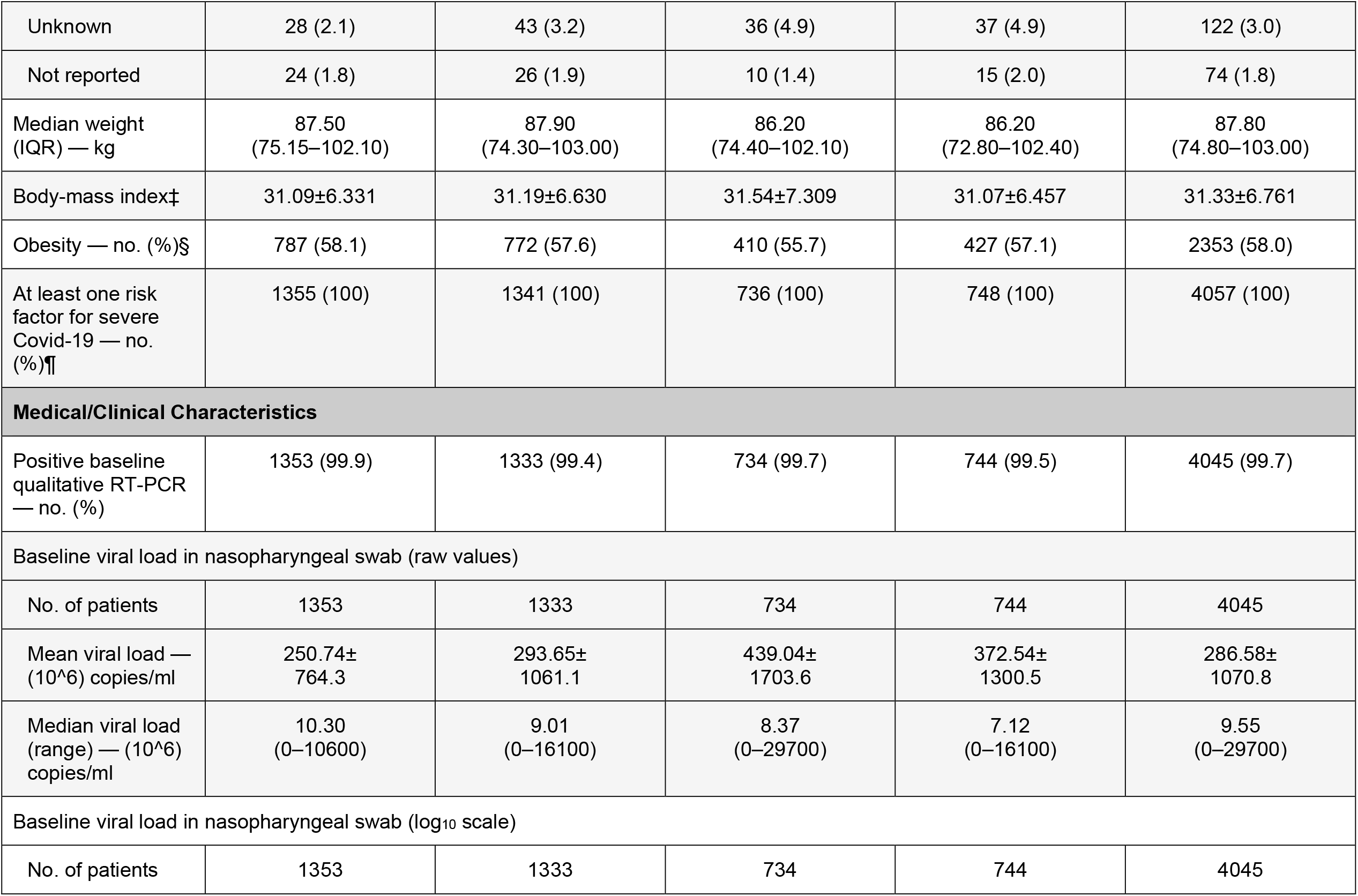
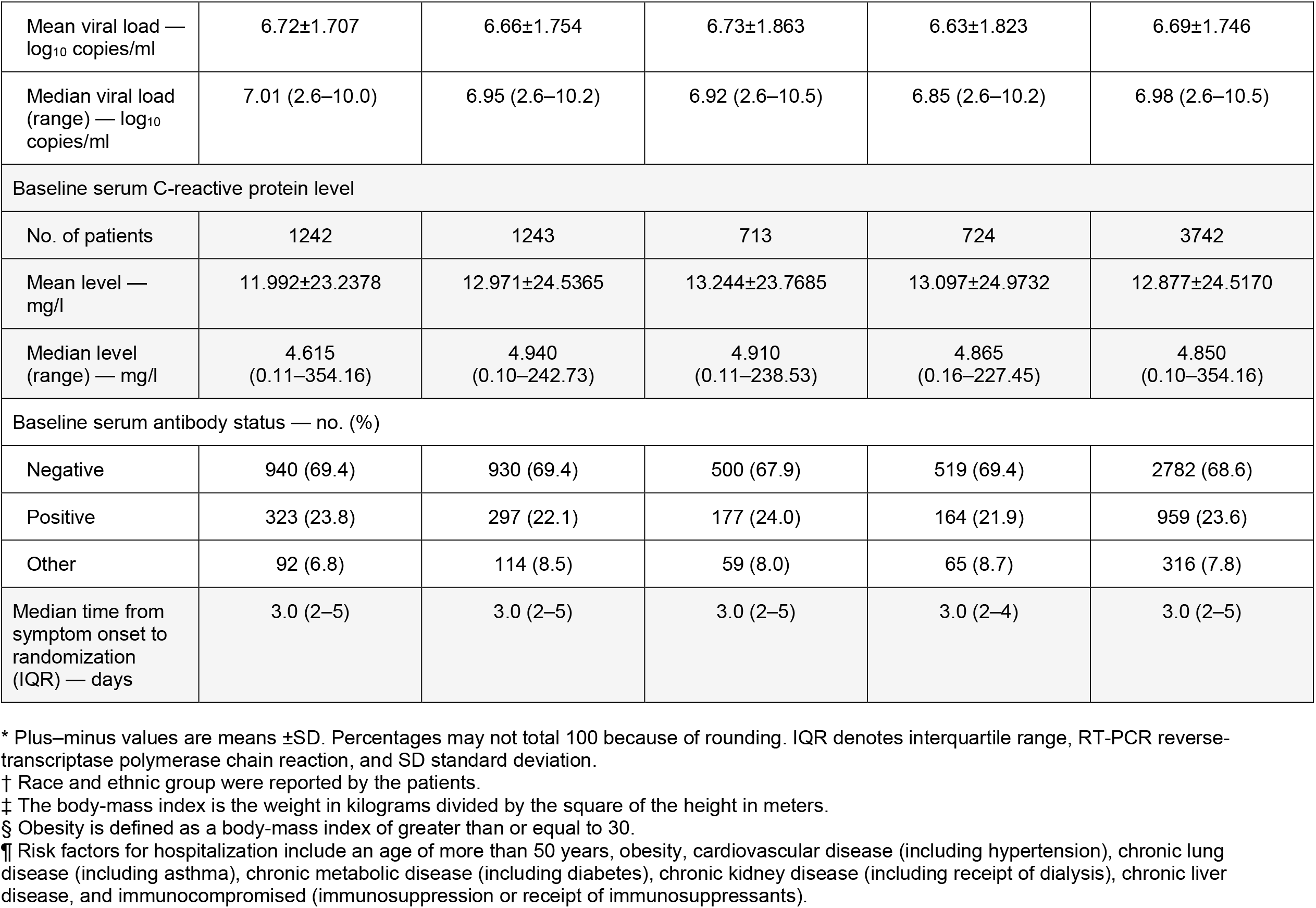
Demographic and Baseline Medical Characteristics* (mFAS)

The median NP viral load was 6.98 log_10_ copies/mL (IQR 5.45-7.85) and the majority of patients (69%) were SARS-CoV-2 serum antibody-negative at baseline (Table 1); these high viral loads and lack of endogenous immune response at baseline suggested that enrolled individuals were early in the course of their infection. Patients had a median of 3 days (IQR 2-5) of Covid-19 symptoms at randomization. NP viral load, serum antibody-negative status, and median days of Covid-19 symptoms at randomization were similar across treatment groups. Demographic and baseline medical characteristics for the REGEN-COV 8000mg mFAS group and concurrent placebo are shown in Table S3.

### NATURAL HISTORY

When considering placebo-treated patients only, there was an association between baseline viral load and Covid-19-related hospitalization or all-cause death: hospitalization/deaths occurred in a greater proportion of patients with high baseline viral load (>10^6^ copies/mL) compared to those with lower viral load (≤10^6^ copies/mL) (6.3% [55/876] vs 1.3% [6/457]; Table S4).

Patients in the placebo group who were serum antibody-negative at baseline had higher median viral loads at baseline compared to those who were serum antibody-positive (7.45 log_10_ copies/ml vs 4.96 log_10_ copies/ml), and took longer to bring their viral levels to below the lower limit of quantification (Figure S2).

Despite these population-level observations, baseline serum antibody status of placebo-treated patients was not predictive of subsequent Covid-19-related hospitalizations or all-cause death, as these rates were similar in patients who were serum antibody-negative and antibody-positive (5.3% [49/930] vs 4.0% [12/297]; Table S4). The lack of positive-predictive value of serum antibody-positive status on the reduction in rates of hospitalization or death suggests that some patients mount an ineffective immune response. For example, placebo patients who were serum antibody-positive but still progressed to requiring hospitalization or died had high viral loads at baseline and day 7, similar to patients who were serum antibody-negative and required hospitalization or died (Table S5).

### EFFICACY

#### Primary Endpoint

REGEN-COV 2400mg and 1200mg similarly reduced Covid-19-related hospitalization or all-cause death by 71.3% (1.3% vs 4.6% placebo; 95% CI: 51.7%, 82.9%; p<0.0001) and 70.4% (1.0% vs 3.2% placebo; 95% CI: 31.6%, 87.1%; p<0.0024), respectively (Table 2; Figure 2A-B; Table S6). Similar reductions in Covid-19-related hospitalization or all-cause death were observed across subgroups, including in patients who were serum antibody-positive at baseline (Table 2; Figure S3).

**Table 2.** Hierarchical End Points.

| Hypothesis Testing Hierarchy* | Comparison | Treatment Effect | P value |
| --- | --- | --- | --- |
| 1. Proportion of patients with $\geq 1$ Covid-19-related hospitalization or all-cause death through day 29 | 2400 mg vs placebo | 18/1355 (1.3%) vs 62/1341 (4.6%);<br>71.3% reduction (95% CI: 51.7%, 82.9%) | <0.0001 |
| 2. Proportion of patients with $\geq 1$ Covid-19-related hospitalization or all-cause death through day 29 | 1200 mg vs placebo | 7/736 (1.0%) vs 24/748 (3.2%);<br>70.4% reduction (95% CI: 31.6%, 87.1%) | 0.0024 |
| 3. Proportion of patients with $\geq 1$ Covid-19-related hospitalization or all-cause death through day 29 in patients with baseline viral load $>10^6$ copies/mL | 2400 mg vs placebo | 13/924 (1.4%) vs 55/876 (6.3%);<br>77.6% reduction (95% CI: 59.3%, 87.7%) | <0.0001 |
| 4. Proportion of patients with $\geq 1$ Covid-19-related hospitalization or all-cause death through day 29 in patients who are serum antibody negative at baseline | 2400 mg vs placebo | 12/940 (1.3%) vs 49/930 (5.3%);<br>75.8% reduction (95% CI: 54.7%, 87.0%) | <0.0001 |
| 5. Proportion of patients with $\geq 1$ Covid-19-related hospitalization or all-cause death through day 29 in patients with baseline viral load $>10^6$ copies/mL | 1200 mg vs placebo | 6/482 (1.2%) vs 20/471 (4.2%);<br>70.7% reduction (95% CI: 27.6%, 88.1%) | 0.0045 |
| 6. Proportion of patients with $\geq 1$ Covid-19-related hospitalization or all-cause death through day 29 in patients who are serum antibody negative at baseline | 1200 mg vs placebo | 3/500 (0.6%) vs 18/519 (3.5%);<br>82.7% reduction (95% CI: 41.6%, 94.9%) | 0.0014 |
| 7. Proportion of patients with $\geq 1$ Covid-19-related hospitalization or all-cause death from day 4 through day 29 | 2400 mg vs placebo | 5/1351 (0.4%) vs 46/1340 (3.4%);<br>89.2% reduction (95% CI: 73.0%, 95.7%) | <0.0001 |
| 8. Proportion of patients with $\geq 1$ Covid-19-related hospitalization or all-cause death from day 4 through day 29 | 1200 mg vs placebo | 5/735 (0.7%) vs 18/748 (2.4%);<br>71.7% reduction (95% CI: 24.3%, 89.4%) | 0.0101 |
| 9. Time to Covid-19 symptoms resolution | 2400 mg vs placebo | Median 10 vs 14 days;<br>4-day faster resolution | <0.0001 |
| 10. Time to Covid-19 symptoms resolution | 1200 mg vs placebo | Median 10 vs 14 days;<br>4-day faster resolution | <0.0001 |

\* All analyses in mFAS: all randomized patients with a positive SARS-CoV-2 RT-qPCR test from nasopharyngeal swabs at randomization and $\geq 1$ risk factor for severe COVID-19.
CI, confidence interval; mFAS, modified full analysis set; RT-qPCR, reverse transcriptase quantitative polymerase chain reaction.

**Figure 2.**
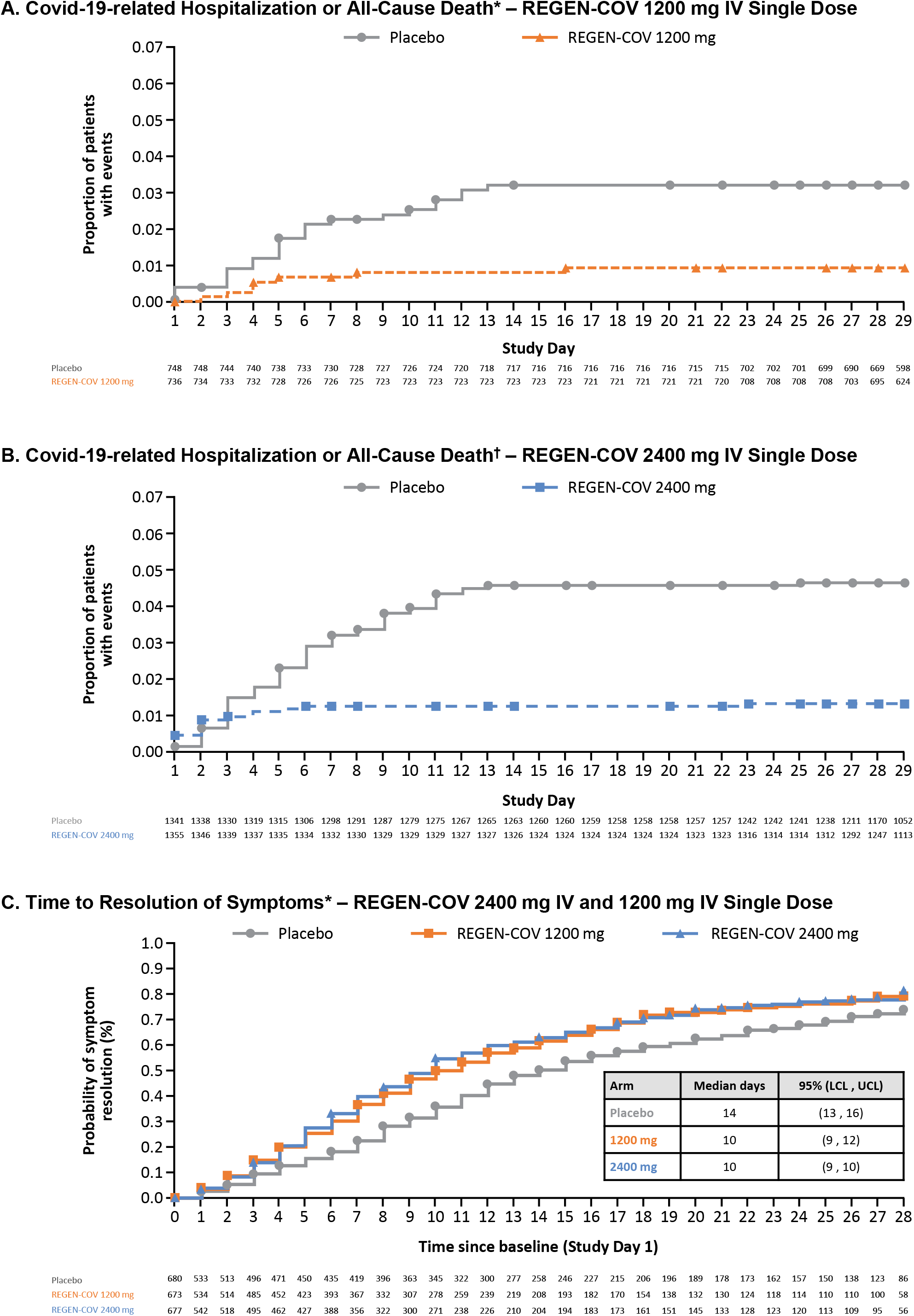
Clinical Efficacy. * In the amended phase 3 portion of the trial. † In the original and amended phase 3 portions of the trial combined. IV, intravenous(ly); LCL, lower confidence limit; UCL, upper confidence limit.

#### Key Secondary Endpoints

The reduction in the proportion of patients with Covid-19-related hospitalization or all-cause death was observed starting approximately 1-3 days after treatment with REGEN-COV (Figure 2A-B). After these first 1-3 days, patients in the placebo group continued to experience Covid-19-related hospitalization or death events during the study period (46/1340 [3.4%]), while very few events occurred in the 2400mg or 1200mg REGEN-COV treatment groups (5/1351 [0.4%] and 5/735 [0.7%], respectively) (Table 2; Figure S4).

The median time to resolution of Covid-19 symptoms was 4 days earlier in both REGEN-COV dose groups than in the placebo groups (10 days vs 14 days, respectively; p<0.0001 each for 2400mg and 1200mg) (Table 2; Figure 2C). The more rapid resolution of Covid-19 symptoms with either dose of REGEN-COV was evident by day 3. Both REGEN-COV doses were associated with similar improvements in symptoms resolution across subgroups (Figure S5).

All REGEN-COV dose levels (1200mg, 2400mg, and 8000mg) led to similar and rapid declines in viral load compared to placebo (Figure S6-S8).

#### Other Secondary Endpoints

REGEN-COV treatment was associated with a lower proportion of patients with Covid-19-related hospitalization (Table S7). Among patients that were hospitalized due to Covid-19, those in the REGEN-COV group had shorter hospital stays and a lower rate of admission to an intensive care unit (Table S8).

REGEN-COV treatment was associated with a lower proportion of patients with Covid-19-related hospitalization, emergency room visits, or all-cause death through Day 29 (Table S9) and a lower proportion requiring any medically-attended visit for worsening Covid-19 (hospitalization, emergency room visit, urgent care visit or physician office/telemedicine visit) or all-cause death (Table S7; Table S10-S11).

### SAFETY

SAEs were experienced by more patients in the placebo group (4.0%) compared to the REGEN COV dose groups (1.1% to 1.7%) (Table 3). More patients experienced TEAEs that resulted in death in the placebo group (5 patients, 0.3%) compared to the REGEN-COV dose groups: 1 (0.1%) in 1200mg, 1 (<0.1%) in 2400mg, and 0 in 8000mg (Table 3; Table S12). Most adverse events were consistent with complications of Covid-19 (Table S13) and the majority were not considered to be related to study drug. Few patients experienced grade ≥2 infusion-related reactions (0 in placebo; 2, 1, and 3 patients in 1200mg, 2400mg, and 8000mg, respectively) and hypersensitivity reactions (1 in placebo and 1 in 2400mg) (Table 3). A similar safety profile was observed between REGEN-COV doses, with no discernable imbalance in safety events.

**Table 3.** Overview of Serious Adverse Events and Adverse Events of Special Interest in the Safety Population.

| Event | Placebo<br>(n=1843) | REGEN-COV<br>1200 mg IV<br>(n=827) | REGEN-COV<br>2400 mg IV<br>(n=1849) | REGEN-COV<br>8000 mg IV<br>(n=1012) | Total<br>(n=5531) |
| --- | --- | --- | --- | --- | --- |
| *no. of patients (percent)* |  |  |  |  |  |
| Serious adverse event that occurred or worsened during the observation period† |  |  |  |  |  |
| Any serious adverse event | 74 (4.0) | 9 (1.1) | 24 (1.3) | 17 (1.7) | 124 (2.2) |
| Any serious adverse event of special interest* | 6 (0.3) | 1 (0.1) | 1 (<0.1) | 1 (<0.1) | 9 (0.2) |
| Adverse events of special interest that occurred or worsened during the observation period† |  |  |  |  |  |
| Grade ≥2 infusion-related reaction within 4 days | 0 | 2 (0.2) | 1 (<0.1) | 3 (0.3) | 6 (0.1) |
| Grade ≥2 hypersensitivity reaction within 29 days | 1 (<0.1) | 0 | 1 (<0.1) | 0 | 2 (<0.1) |
| Patients requiring medical attention related to Covid-19 at a healthcare facility | 47 (2.6) | 15 (1.8) | 20 (1.1) | 11 (1.1) | 93 (1.7) |
| Patients requiring medical attention not related to Covid-19 at a healthcare facility | 5 (0.3) | 0 | 7 (0.4) | 0 | 12 (0.2) |
| Adverse events that occurred or worsened during the observation period† |  |  |  |  |  |
| Patients with event | 189 (10.3) | 59 (7.1) | 142 (7.7) | 85 (8.4) | 475 (8.6) |
| Patients with grade 3 or 4 event | 62 (3.4) | 11 (1.3) | 18 (1.0) | 15 (1.5) | 106 (1.9) |
| Patients with event that led to death | 5 (0.3) | 1 (0.1) | 1 (<0.1) | 0 | 7 (0.1) |
| Patients with event that led to withdrawal from the trial | 1 (<0.1) | 0 | 1 (<0.1) | 2 (0.2) | 4 (<0.1) |
| Patients with event that led to infusion interruption* | 0 | 1 (0.1) | 0 | 1 (<0.1) | 2 (<0.1) |

\* Events were hypersensitivity reactions (grade $\geq 2$ ), infusion-related reactions (grade $\geq 2$ ), or medical attention at a healthcare facility, regardless of relation to Covid-19.
† Events listed here were not present at baseline or were an exacerbation of a preexisting condition that occurred during the observation period, which is defined as the time from administration of REGEN-COV or placebo to the final follow-up visit.

### PHARMACOKINETICS

The mean concentrations of casirivimab and imdevimab in serum on day 29 increased in a dose-proportional manner and were consistent with linear pharmacokinetics (Table S14). The mean day 29 concentrations of casirivimab and imdevimab in serum were 46.4±SD22.5 and 38.3±SD19.6 mg/L, respectively, for the 1200mg dose and 73.2±SD27.2 and 60.0±SD22.9 mg/L, respectively, for the 2400mg dose; the mean estimated half-life was 28.8 days for casirivimab and 25.5 days for imdevimab (Table S14).

## DISCUSSION

Previous Phase 1/2 data showed that, in outpatients with Covid-19, REGEN-COV lowered viral load, reduced the need for medical attention, and was highly suggestive of a reduced risk for hospitalization.^16^ The phase 3 clinical outcomes data presented here confirm that early treatment with REGEN-COV in outpatients with risk factors for severe Covid-19 can dramatically lower the risk of hospitalization or all-cause death. Both 1200mg IV and 2400mg IV REGEN-COV led to a ≥70% reduction (vs placebo) in Covid-19 hospitalization or all-cause death over 28 days after treatment. In those who were hospitalized, REGEN-COV treatment led to shorter duration of hospitalization and a lower proportion of patients requiring intensive care unit (ICU)-level care. In addition, REGEN-COV, at both doses, resulted in more rapid resolution of Covid-19 symptoms by a median of 4 days. Therefore, a single dose of REGEN-COV in outpatients with Covid-19 has the potential to improve patient outcomes and substantially reduce the health care burden experienced during this pandemic by reducing morbidity and mortality, including hospitalizations and ICU-level care. Furthermore, REGEN-COV can substantially speed up recovery from Covid-19, which represents an additional benefit for patients as there is a growing body of evidence that some patients, including those with mild symptoms, will have a variably prolonged course of recovery.^18-20^

We previously hypothesized that, while host factors play a role in the disease course, the morbidity and mortality of SARS-CoV-2 result from high viral burden such that early treatment with an anti-spike monoclonal antibody combination could reduce this risk. In the placebo group, we found that patients with hospitalizations or all-cause death had markedly higher viral loads at baseline and were slower to clear virus, independent of baseline serological status. Patients in the placebo group who had mounted their own endogenous antibody response to SARS-CoV-2 (serum antibody-positive) had similar rates of hospitalizations or death compared to patients who were serum antibody-negative, suggesting that some serum antibody-positive patients had an ineffective immune response. Furthermore, placebo patients who were serum antibody-positive and had a Covid-19-related hospitalization or who died, also had high baseline viral load levels similar to patients who were serum antibody-negative and had these events, supporting high viral load as the key driver of severe Covid-19. Moreover, this study demonstrated that there is clinical benefit of REGEN-COV, regardless of baseline serum antibody status, making serological testing at the time of Covid-19 diagnosis less critical for clinical treatment decisions. This is important given the prevalence of vaccine utilization, which will result in baseline serum antibody-positive status that may not effectively prevent severe infection in patients with ineffective natural immunity (as appears to be the case for certain patients in this trial) or in the setting of emerging VOC/VOIs.

Both 1200mg and 2400mg doses of REGEN-COV had similar antiviral and clinical efficacy, suggesting that, in this study, drug concentrations were well above the minimally effective dose. Both doses rapidly reduced viral loads with faster time to viral clearance compared to placebo. In addition to providing clinical benefit to the individual patient receiving REGEN-COV, the rapid antiviral effect is likely to be associated with a public health benefit through reduced risk of viral transmission and containment of SARS-CoV-2 VOC/VOIs.

A low incidence of serious adverse events and hypersensitivity and infusion-related reactions was observed. Similar to results reported previously,^16^ concentrations of each antibody in serum through day 29 were well above the predicted neutralization target concentration based on in vitro and preclinical data.

The emergence of resistant variants of SARS-CoV-2 during treatment with an antiviral agent(s) or via circulation within the global community will continue to be a challenge for the success of Covid-19 therapeutics and vaccines. Although in vitro studies or in vivo animal studies demonstrate that combinations of non-competing antibodies, such as REGEN-COV, are able to suppress the emergence of resistant variants, the relevance of those studies to natural human infection is uncertain. We therefore recently investigated and reported the genetic diversity of the spike protein across samples from 1,000 REGEN-COV- or placebo-treated patients enrolled into either this outpatient trial (NCT04425629) or a separate, hospitalized Covid-19 trial (NCT04426695). The analysis of 4,882 samples from these 1,000 patients demonstrated that REGEN-COV protects against the selection of resistant variants, as evidenced by a similar number of receptor binding domain (RBD) variants found in placebo-treated (15 variants) and REGEN-COV-treated (12 variants each in 1200mg and 2400mg groups) patients.^14^

REGEN-COV at the 2400mg dose received Emergency Use Authorization (EUA) from the US FDA in November 2020 for the treatment of mild-to-moderate Covid-19.^15,21^ On April 8, 2021, the NIH treatment guidelines recommended the use of 2400mg REGEN-COV for the treatment of high-risk outpatients with Covid-19, with preferential use of REGEN-COV in areas where VOC/VOIs are common.^22^ The clinical evidence from this clinical outcomes trial, the largest randomized, controlled phase 3 Covid-19 outpatient treatment trial to date, indicates that, similar to the 2400mg dose, 1200mg REGEN-COV is well-tolerated, can significantly reduce Covid-19-related hospitalizations or death, can speed time to recovery, and is unlikely to promote the emergence of treatment-resistant SARS-CoV-2 variants. These definitive phase 3 data demonstrate a profound reduction in the risk of hospitalization or all-cause death, together with an acceptable safety profile, in high-risk, SARS-CoV-2-positive adults. The results support wide access of this treatment modality so as to lower the morbidity and mortality associated with SARS-CoV-2 infection.

## Supporting information

CONSORT Checklist

AM ICMJE

BM ICMJE

CK ICMJE

CP ICMJE

DA ICMJE

DR ICMJE

DW ICMJE

GY ICMJE

HG ICMJE

JB ICMJE

JD ICMJE

JH ICMJE

JI ICMJE

LL ICMJE

LR ICMJE

NB ICMJE

RB ICMJE

SA ICMJE

SS ICMJE

TD ICMJE

WK ICMJE

YK ICMJE

## Data Availability

Qualified researchers may request access to study documents (including the clinical study report, study protocol with any amendments, blank case report form, statistical analysis plan) that support the methods and findings reported in this manuscript. Individual anonymized participant data will be considered for sharing once the indication has been approved by a regulatory body, if there is legal authority to share the data and there is not a reasonable likelihood of participant re-identification. Submit requests to https://vivli.org/.

## DATA SHARING

A data sharing statement provided by the authors is available with the full text of this article.

## SUPPORTED BY

Supported by Regeneron Pharmaceuticals, Inc. Certain aspects of this project have been funded in whole or in part with federal funds from the Department of Health and Human Services; Office of the Assistant Secretary for Preparedness and Response; Biomedical Advanced Research and Development Authority, under OT number: HHSO100201700020C.

## ACKNOWLEDGEMENT

We thank the study participants; their families; the investigational site members involved in this trial (principal and subprincipal investigators, listed in the Supplementary Appendix); the Regeneron trial team (members listed in the Supplementary Appendix); the members of the independent data and safety monitoring committee; Brian Head, Ph.D., Caryn Trbovic, Ph.D., and S. Balachandra Dass, Ph.D., from Regeneron Pharmaceuticals for assistance with development of an earlier version of the manuscript; and Prime for formatting and copy editing suggestions for an earlier version of the manuscript.

