## Supplementary Appendix for "REGEN-COV Antibody Cocktail Clinical Outcomes Study in Covid-19 Outpatients"

#### Study Sites and Investigators

**<u>Advanced Pulmonary Research Institute, Loxahatchee, FL</u>:** Neal Warshoff, Liudmila Moreiras

**<u>AGA Clinical Trials, Miami, FL</u>:** Dario Altamirano, Dickson Ellington, Faisal Fakih

**<u>Arizona Liver Health, Tucson, AZ</u>:** Anita Kohli, Vicki McIntyre, Yessica Sachdeva, Ashley Carney

**<u>Arizona Liver Health, Puyallup, WA:</u>** Yessica Sachdeva, Anita Kohli, Amanda McFarland, Dina Gibson, Victorine Ekoko

**<u>Ark Clinical Research, Long Beach, CA</u>:** Kenneth Kim, Lisa Neinchel, Nayna Paryani, Amber Mottola, Eva Day, Martha Navarro, Apinya Vutikullird

**<u>Atella Clinical Research La Palma, CA</u>:** Rafaelito Victoria, Xanthe Victoria, Rene Uong

**<u>Atrium Health, Charlotte, NC</u>:** Christopher Polk, Mindy Sampson, Michael Leonard, Lewis McCurdy, Leigh Ann Medaris, Zainab Shahid, Lisa Davidson

**<u>Avera McKennan Hospital and University Health Center, Sioux Falls, SD</u>:** Jawad Nazir, John Lee, Amy Elliott, Touba Naim, Khizar Hamid, Muhammad Hamza, Robert Kessler, Kara Bruning

**<u>Axces Research, Sante Fe, NM</u>:** Linda Gorgos, Erika Benson, Michael Palestine

**<u>Bio-Medical Research, LLC, Miami, FL</u>:** Lilia Roque-Guerrero, Ana Gomez Ramirez, Javier Capote, Gisel Paz

**<u>Carolina Medical Research, Clinton, SC</u>:** Nancy Patel, Ravikumar Patel, Ryan Sattar

**<u>Catalina Research Institute, Montclair, CA</u>:** Rizwana Mohensi, Shelia De Jesus-Maranan, Cecilia Casaclang

**<u>Centex Studies, Lake Charles, LA</u>:** Michael Seep, Celeste Brown, Joshua Whatley

**<u>Chicago Clinical Research Institute, Chicago, IL</u>:** Dennis Levinson, Norman James, Saad Alvi, Azazuddin Ahmed

**Clinical Research of Central Florida, Winter Haven, FL:** Robinson Koilpillai, Stephanie Cassady, Jennifer Cox, Eduardo Torres

**Crossroads Clinical Research, Corpus Christi, TX:** Michael Winnie, Jerry Plemons, Omesh Verma, Richard Leggett

**DM Clinical Research/BFHC Research, San Antonio, TX:** Ramon Reyes, Keith Beck, Brian Poliquin

**DM Clinical Research/LinQ Research, LLC, Pearland, TX:** Murtaza Mussaji, Jignesh Shah

**Duke Clinical Research, Durham, NC:** John Eppensteiner, Alexander Limkakeng, Joseph Borawski, Samuel Francis, Charles Gerardo, Emily Thatcher, Harajeshwar Kohli, Rachel O’Brian

**EME RED Hospitalaria, Mérida, Mexico:** Rodrigo Buendia, Natalia Romero Pavía, Juan Francisco Rubio Suárez, Mario Humberto Bustillos Pech

**Epic Medical Research, Red Oak, TX:** Haresh Boghara, Sunny Patel, Bari Eichelbaum

**Eukarya Pharmasite S.C., Nuevo Leon, Mexico:** Ricardo Tellez, Stephani Moreno

**Excel Clinical Research, Las Vegas, NV:** Duane Anderson, Sean Su, Alexander Akhavan, Diana Kirby, Joy Venglik, Crista Fedora

**FAICIC S. de RL de C.V., Veracruz, Mexico:** Alejandro Quintín Barrat Hernández, Silvano Omar Martinez Pérez, Edgar Iván Muñoz López

**Florida Pulmonary Research Institute, LLC, Winter Park, FL:** Faisal A. Fakih, Faisal M. Fakih, Fernando Alvarado, Daniel Layish, Jose Diaz, Andres Perez

**Fred Hutchinson Cancer Research Centre, Seattle, WA:** Michelle Karuna, Michael Boeckh, Elizabeth Church, Alison Roxby,

**Future Innovative Treatments, LLC, Colorado Springs, CO:** Bhaktasharan Patel, Gary Tarshis, Katrina Grablin

**Global Clinical Professionals Research, St Petersburg, FL:** Roxana Stoici, Gualberto Perez, Joseph Pica, Enrique Villareal

**Harlem Hospital Center – NYCHHC, Harlem, NY:** Farbod Raiszadeh, Sharon Mannheimer, Khaing Myint, Hussein Assallum, Lovelyamma Varghese, Akari Kyawa

**Holy Name Medical Center, Teaneck, NJ:** Suraj Saggar, Thomas Birch, Benjamin De La Rosa, Karyna Neyra, Erina Kunwar

**Hope Clinical Research, Canoga Park, CA:** Hessam Aazami, Cheryl Bland, Deborah Wu, Jamsheed Akhavan

**Hospital Angeles Chihuahua, Chihuahua, Mexico:** Belinda Sofia Gomez Quintana, Hector Rascón Marquez

**Houston Methodist Hospital, Houston, TX:** Howard Huang, Jihad Georges Youssef, Simon Yau, Ahmad Goodarzi, Mukhtar Al-Saadi, Faisal Zahiruddin

**IACT Health, Columbus, GA:** Jeffrey Kingsley, April Pixler

**Icahn School of Medicine at Mount Sinai, Manhattan, NY:** Judith Aberg, Michelle Cespedes, Alexandra Abrams-Downey, Erna Kojic, Luz Lugo, Sean Liu, Nadim Salomon, David Perlman, Deena Altman, Farah Rahman, Georgina Osorio, Joseph Mathew, Sanjana Koshy, Dana Mazo, Francesca Cossarini, Sondra Middleton, Alina Jen, Erika Maria Reategui Schwarz

**Innova Health Care Services (INOVA Fairfax Hospital), Falls Church, VA:** Christopher deFilippi, Steven Nathan, Lindsay Clevenger

**Instituto de Investigaciones Clínicas para la Salud A.C. Durango, Mexico:** Isabel Buendia Suarez, Cristina Resendez

**Jacobi Medical Center, Bronx, NY:** Gabriele DeVos, David Stein, Jason Leider, Kellie Roe, Jane Devereux, Elizabeth Jenny-Avital

**Köhler and Milstein Research SA de CV, Merida, Yucatan, Mexico:** Jesus Abraham Simon Campos, Felipe de Jesús Pineda Cárdenas

**Lincoln Medical Center – NYCHHC, Bronx, NY:** Vidya Menon, Moiz Kasubhai, Usha Venugopal, Anjana Pillai, Franscene Oulds, Paola Carugno, Daniel Sittler

**Long Beach Medical Center, Long Beach, CA:** Jimmy Johannes, Thomas Jaing, Christopher Yee, Henry Su, Andrew Wittenberg, Anthony Arguija

**Maryland School of Medicine, Baltimore, MD:** Richard Wilkerson, Shyam Kottilil, Shivakumar Narayanan, Joel Chua, Jennifer Husson, John Baddley

**Medical Research of Westchester, Miami, FL:** Richard Perez-Perez, Carlos J. Bello, Esperanza Arce-Nunez, Jorge Acosta, Julio L. Arronte

**Medical University of South Carolina, Charleston, SC:** Eric Meissner, Patrick Flume, Andrew Goodwin, Deeksha Jandhyala, Nandita Nadig

**Mercury Clinical Research, Houston, TX:** Rajasekaran Annamalai, Huy Nguyen, Nizar Nayani, Mahalakshmi Ramchandra

**META Medical Research Institute, Dayton, OH:** Priyesh Mehta, Jacqueline Horne, Grace Hassan

**Miami Valley Hospital, Dayton, OH:** Thomas Herchline, Steve Burdette, Jonathan Pope, David Herman

**Midland Florida Clinical Research Center, Deland, FL:** Godson Oguchi, DeAndrea Duffus

**Midway Immunology and Research Center, Fort Pierce, FL:** Moti Ramgopal, Brenda Jacobs, Lisa Cason, Angela Trodglen

**Next Level Urgent Care, Houston, TX:** Terence Chang, Robbyn Traylor, Lenee Gordon, John McDivitt, Lizette Castro

**Palos Verdes Medical Group (PVMG), Peninsula Research Associates, Inc., Rolling Hills Estates, CA:** Lawrence Sher, Monica Saad, LeighAnn Schmidt

**Pharma Tex Research, LLC, Amarillo, TX:** David Brabham, Mark Sigler, Tarek Naguib

**PMG Research of McFarland Clinic, Ames, IA:** Jennifer Killion, Rupal Amin, Shauna Basener, Timothy Lowry

**PMG Research of Wilmington, Wilmington, NC:** Kevin Cannon, Mesha Chadwick

**Providence Saint John's Health Center, Santa Monica, CA:** Terese Hammond, Fabian Andres Romero, Steven O'Day, Trevan Fischer, Ana Rocha, Anmol Rangoola

**Qway Research, Hialeah, FL:** Oscar Galvez, Fausto Castillo

**Regional One Health, Puyallup, WA and Memphis, TN:** John Jefferies, Scott Strome, Sandy Arnold, Terri Finkel, Amber Thacker, Amik Sodhi, Elisha McCoy, Daniel Wells, Nathaniel Rogers, David Schwartz

**Remington-Davis, Columbus, OH:** Edward Cordasco, Brian Zeno, Heather Holmes, Heather Lee

**Rhode Island Hospital, Providence, RI:** Eleftherios Mylonakis, Dimitrios Farmakiotis, Natasha Ryback, Karen Tashima, Francesca Beaudoin, Selim Suner, Gregory Jay, Katelyn Moretti, Adam Aluisio, Naz Karim, Sonya Naganathan, William Binder, Adam Levine, Neel Belani, John Lee, Taneisha Wilson, Anshul Parulkar, Ramu Kharel, Alexis Lawrence

**RM Pharma Specialists S.A. de Colonia del Valle, Mexico City, Mexico:** Lucero Sanchez, Ana Karla Guzmán Romero

**Ruane Clinical Research Group, Los Angeles, CA:** Peter Ruane, Peter Wolfe, Kenny Trinidad, Isaac Berlin

**San Francisco Research Institute, San Francisco, CA:** Mark Savant, Francis Hsiao, Edna Yee

**Sarasota Memorial Hospital, Sarasota, FL:** Manuel Gordillo, Rishi Bhattacharyya, Sudha Tallapragada, Annette Artau, Julie Larkin, Roberto Mercado, Michael Milam, Natan Kraitman, Sarah Temple, Lenka Offner, Rabih Loutfi, Kirk Voelker, Michael Lowry, Marshall Frank, Ashley Grant

**SignatureCare Emergency Center – TC Jester, Houston, TX:** Alan Skolnick, Harold Minkowitz, David Leiman, Todd Price, Anatoli Krasko

**Stanford University, Palo Alto, CA:** Upinder Singh, Aruna Subramanian, Yvonne Maldonado, Jason Andrews, Chaitan Khosla

**Sun Research Institute, San Antonio, TX:** Carl Dukes, Robert Bass, Larry Lothringer, Leonel Reyes

**Tandem Clinical Research, Maitland, FL:** Esteban Olivera, Mayra Abreu

**<u>Tandem Clinical Research, Marrero, LA</u>:** Adil Fatakia, Marissa Miller, Kristen Clinton, Gary Reiss

**<u>Temple University Hospital (TUH), Philadelphia, PA</u>:** Gerard Criner, Nathaniel Marchetti, Parag Desai, Daniel Salerno, Fredric Jaffe, Samuel Krachman, Matthew Zheng, Maulin Patel, Junad Chowdhury, Daniel Mueller

**<u>The George Washington University Hospital, Washington, WA</u>:** David Diemert, Afsoon Roberts, David Parenti, Hana Akselrod, Marc Siegel, Andrew Meltzer, Elissa Malkin, Gary Simon

**<u>Triple O Research Institute PA, West Palm Beach, FL</u>:** Olayemi Osiyemi, Jose A. Menajovsky-Chaves, Christina Campbell

**<u>Tulane University School of Medicine, New Orleans, LA</u>:** Dahlene Fusco, Arnaud Drouin, Joshua Denson, Jerry Zifodya, Christine Bojanowski, Monika Dietrich, Stacy Drury

**<u>Universal Medical and Research Center, LLC, Miami, FL</u>:** Gerard Acloque**,** Agustin Martinez

**<u>University of California (UC) Davis, Sacramento, CA</u>:** Timothy Albertson, Nicholas Kenyon, Brian Morrissey, Christian Sandrock, Stuart Cohen

**<u>University of Iowa, Iowa City, IA</u>:** Alejandro Comellas, Joel Kline, Spyridon Fortis

**<u>University of Mississippi Medical Center, Jackson, MS</u>:** Gailen Marshall, Utsav Nandi, Vishnu Garla, John Spurzem, Andrew Wilhelm

**<u>University of South Florida, Tampa, FL</u>:** Kami Kim, Seetha Lakshmi, Tiffany Vasey, Asa Oxner, Jason Wilson, Lucy Guerra

**<u>University of Texas (UT) – Southwestern Medical Center, Dallas, TX</u>:** Satish Mocherla, Mamta Jain, Jessica Meisner, Nancy Rollins

**<u>Wellstar Kennestone Hospital, Marietta, GA</u>:** Danny Branstetter, Neha Paranjape

**<u>Willis-Knighton Physician Network, Shreveport, LA</u>:** Joseph Bocchini, Clint Wilson

**<u>Xera Med Research, Boca Raton, FL</u>:** Anna Martin, Gargi Gharat, Candace Kokaram, Ket Wray, Clement Partap, Ulyana Arzamasova, Kristina Louissaint, Maria Fernandez

**<u>Xera Med Research, Miami, FL</u>:** Anna Martin, Ket Wray, Kristina Louissaint, Maria Fernandez, Gargi Gharat

#### Regeneron Study Team

Achint Chani, Adebiyi Adepoju, Adnan Mahmood, Aisha Mortagy, Ajla Dupljak, Alexander Kansky, Alison Brown, Alpana Waldron, Amanda Cook, Amy Froment, Andrea Hooper, Andrea Margiotta, Andrew Bombardier, Anne Smith, Aswani Bathula, Bari Kowal, Barry Siliverstein, Benjamin Horel, Bret Musser, Brian Bush, Brian Head, Bryan Zhu, Camille Debray, Careta Phillips, Carol Lee, Caryn Trbovic, Catherine Elliott, Chad Fish, Charlie Ni, Charlotte Lyon, Christina Perry, Christine Enciso, Christopher Caira, Christopher Chamak, Christopher Powell, Cliff Baum, Colby Burk, Crystal LaPoint, Cynthia Pan, Danise Subramaniam, David Liu, David Stein, Daya Gulabani, Deborah Leonard, Denise Bonhomme, Denise Kennedy, Derrick Bramble, Dhanalakshmi Barron, Diana Rofail, Dipinder Kaur, Dominique Atmodjo Watkins, Dona Bianco, Donna Gambaccini, Eduardo Forleo Neto, Edward Jean-Baptiste, Ehsan Bukhari, Elizabeth Bucknam, Emily Nanna, Esther Huffman O'Keefe, Evelyn Gasparino, Georgia Bellingham, Giane Sumner, Grainne Moggan, Grainne Power, Haitao Gao, Haixia Zeng, Hannah Smith, Heath Gonzalez, Helen Kang, Hibo Noor, Ian Minns, James Donohue, Janice Austin, Janie Parrino, Jeannie Yo, Jenna McDonnell, Jennifer Hamilton, Jessica Boarder, Jing Xiao, Jingchun Yu, Joanne Malia, Joanne Tucciarone, John Strein, Jonathan Cohen, Jordan Ursino, Joseph Im, Joseph Wolken, Karen Browning, Karen Yau, Kenneth Turner, Kimberly Dornheim, Kit Chiu, Kristina McGuire, Kristy Macci, Kurt Ringleben, Kyle Foster, Lacey Douthat, Laura Sarkis, Linda Kelly, Latora Knighton, Lisa Boersma, Lisa Hersh, Lisa Purcell, Lisa Sherpinsky, Lori Geissler, Mabel Osa-Joachimo, Martha Simpkins, Nagaratna Reddy Medapti, Nagendher Burra, Naresh Lall, Neena Sarkar, Nicholas Moore, Nicole Memblatt, Nikki Miocevic, Nirav

Shah, Nitin Kumar, Nkechi Moghalu, Pallavi Rajput, Patricia Humphries, Pradeep Thanigaimani, Purushottam Risal, Rafia Bhore, Sara Dale, Sonia Yanes, Steven Chen, Suzanne Luther, Yuming Zhao

### Supplementary Methods

#### Symptoms Evolution of COVID-19 (SE-C19)

The Symptoms Evolution of COVID-19 (SE-C19) instrument was an electronic diary that was completed daily from Day 1 to Day 29. The SE-C19 was initially developed based on the CDC symptom list and available published literature specific to patients with COVID-19. It included a list of 23 symptoms feverish, chills, sore throat, cough, shortness of breath or difficulty breathing, nausea, vomiting, diarrhea, headache, red or watery eyes, body aches, loss of taste or smell, fatigue, loss of appetite, confusion, dizziness, pressure or tight chest, chest pain, stomachache, rash, sneezing, sputum or phlegm, runny nose). Patients indicated which of the 23 symptoms they experienced in the last 24 hours and then rated each symptom selected at its worst moment in that period on a scale of mild, moderate or severe.

In parallel to the main clinical trial, patient and clinician interviews were performed to confirm the content validity of the newly developed SE-C19 and psychometric validation was conducted using blinded phase 1/2 data to explore the reliability and validity of the measure and refine a symptom endpoint. The results indicated 19 of the original 23 items being most valid, reliable and relevant to outpatients with COVID-19 (i.e., sneezing, rash, vomiting and confusion were excluded) and refinement of the response options to three-categories (0 – none, 1 – mild/moderate, 2 – severe). The detailed, rigorous scientific methods implemented and results of these additional studies will be published independently.

#### Missing Data Handling

Missing data for virology endpoints was handled as follows: Analysis-positive polymerase chain reaction (PCR) results below the lower limit of quantification (LLOQ) of 714 copies/ml (2.85 log10 copies/ml) were imputed as half the LLOQ (357 copies/ml) and negative PCR results were imputed as 0 $\log_{10}$ copies/ml (1 copy/ml).

Patients with missing baseline symptom assessment were not included in the analysis of the symptom resolution endpoint. Patients who do not experience resolution of symptoms will be censored at the last observation time point up to day 29. Patients who died or had COVID-19-related hospitalization prior to day 29 were censored at day 29.

#### Measurement of REGN10933 and REGN10987 in Serum

Prior to protocol amendment 6, serum for drug concentration analysis was collected from all patients randomized to 2.4 g IV, 8.0 g IV, or placebo at pre-dose (at the screening or baseline visit), day 1 at the end of the infusion, and day 29. After protocol amendment 6, serum for drug concentration analysis was collected from patients randomized to 1.2g IV, 2.4g IV, or placebo in a PK sub-study at pre-dose (at the screening or baseline visit), day 29, and day 120.

The human serum concentrations of REGN10933 (casirivimab) and REGN10987 (imdevimab) were measured using validated immunoassays which employ streptavidin microplates from Meso Scale Discovery (MSD, Gaithersburg, MD, USA). The methods utilized two anti-idiotypic monoclonal antibodies, each specific for either REGN10933 or REGN10987, as the capture antibodies. Captured REGN10933 and REGN10987 were detected using two different, non-competing anti-idiotypic monoclonal antibodies, each also specific for either REGN10933 or REGN10987. The bioanalytical methods specifically quantitated the levels of each anti-SARS-CoV-2 spike monoclonal antibody separately, with no interference from the other antibody. The assay has an LLOQ of 0.156 μg/ml for each analyte in the undiluted serum sample.

### Supplementary Figures

#### Figure S1. Schematic Overview of the Study Design

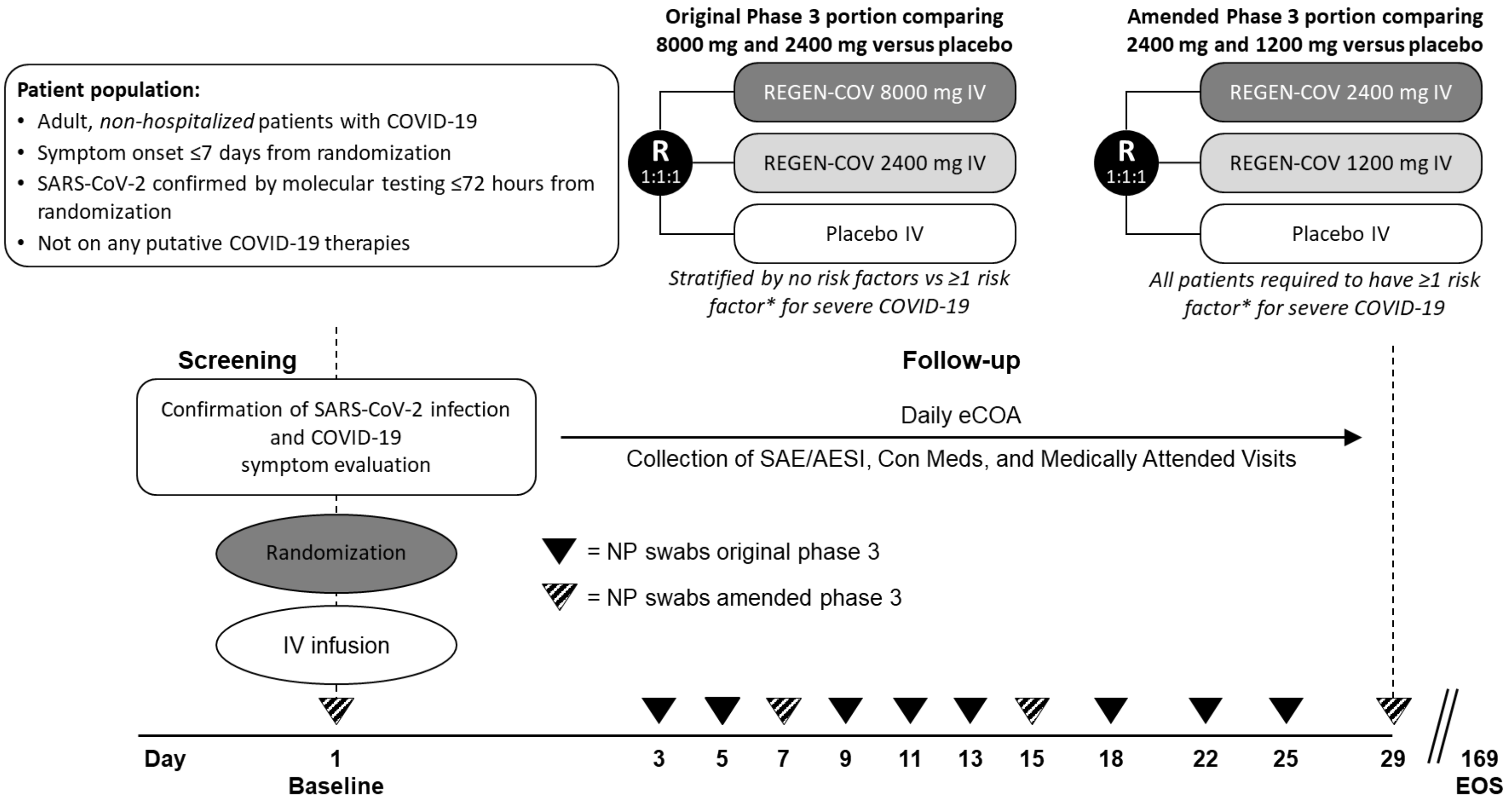

* Risk factors were defined as age ≥50 years, obesity (BMI >30 kg/m$^2$), cardiovascular disease, including hypertension, type 1 or 2 diabetes mellitus, chronic lung disease, including asthma, chronic liver disease, chronic kidney disease, and immunocompromised.

AESI, adverse event of special interest; con med, concomitant medication; eCOA, electronic clinical outcome assessment; EOS, end of study; IV, intravenous(ly); NP, nasopharyngeal; R, randomized; SAE, serious adverse event.

#### Figure S2. Viral Load Over Time in the Placebo Arm by Baseline Serum Antibody Status

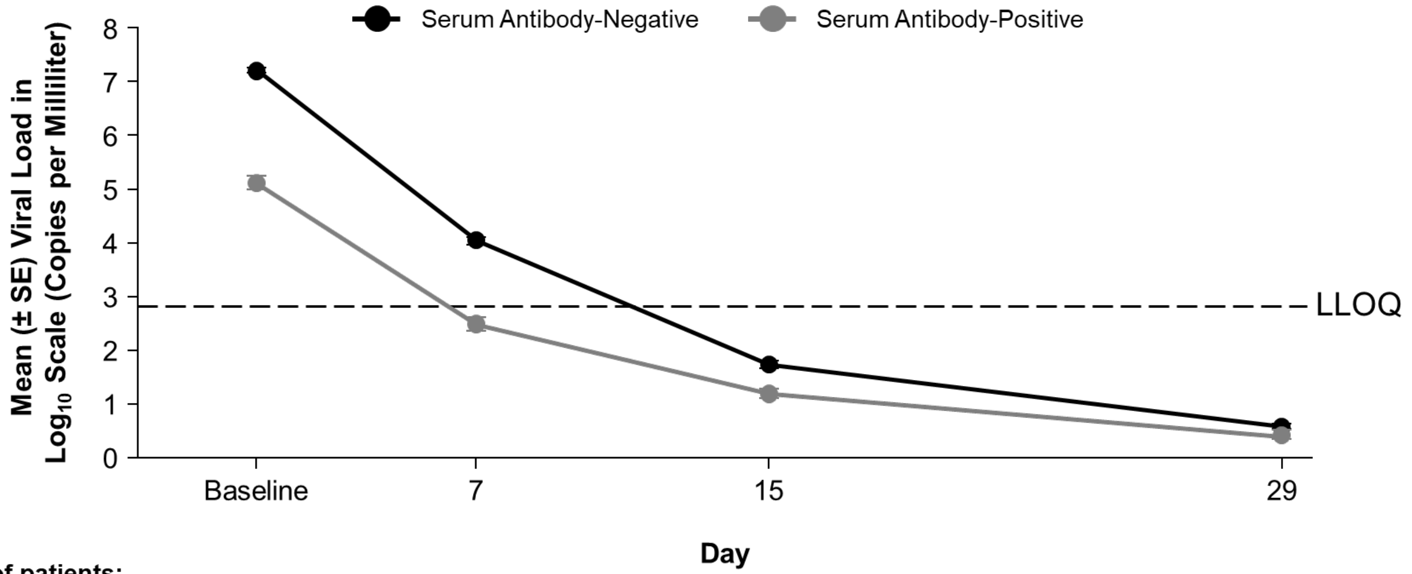

**Number of patients:**

| | | | | |
|---|---|---|---|---|
| Serum Antibody-Negative | 926 | 857 | 836 | 846 |
| Serum Antibody-Positive | 295 | 275 | 265 | 261 |

LLOQ, lower limit of quantification; SE, standard error.

#### Figure S3. Forest Plots: COVID-19-related Hospitalization or All-Cause Death Through Day 29

##### A. Subgroups: Baseline viral load and serum antibody status

| | REGEN-COV Events/N (%) | Placebo Events/N (%) | Rel. Risk Reduction (95% CI) | P |
|---|---|---|---|---|
| **mFAS** | | | | |
| 1200 mg vs Placebo | 7/736 (1.0) | 24/748 (3.2) | 70% (32%, 87%) | 0.0024 |
| 2400 mg vs Placebo | 18/1355 (1.3) | 62/1341 (4.6) | 71% (52%, 83%) | <0.0001 |
| **Baseline Viral Load >10^6 copies/mL** | | | | |
| 1200 mg vs Placebo | 6/482 (1.2) | 20/471 (4.2) | 71% (28%, 88%) | 0.0045 |
| 2400 mg vs Placebo | 13/924 (1.4) | 55/876 (6.3) | 78% (59%, 88%) | <0.0001 |
| **Baseline Viral Load <=10^6 copies/mL** | | | | |
| 1200 mg vs Placebo | 1/252 (0.4) | 4/273 (1.5) | 73% (-141%, 97%) | 0.3746 |
| 2400 mg vs Placebo | 5/429 (1.2) | 6/457 (1.3) | 11% (-189%, 73%) | 1 |
| **Baseline Seronegative** | | | | |
| 1200 mg vs Placebo | 3/500 (0.6) | 18/519 (3.5) | 83% (42%, 95%) | 0.0014 |
| 2400 mg vs Placebo | 12/940 (1.3) | 49/930 (5.3) | 76% (55%, 87%) | <0.0001 |
| **Baseline Seropositive** | | | | |
| 1200 mg vs Placebo | 1/177 (0.6) | 6/164 (3.7) | 85% (NA, 98%) | 0.0588 |
| 2400 mg vs Placebo | 4/323 (1.2) | 12/297 (4.0) | 69% (6%, 90%) | 0.04 |

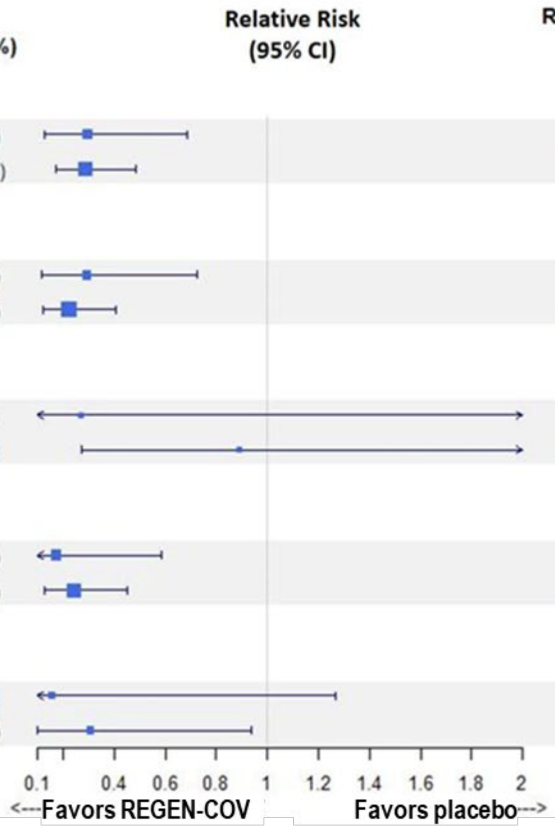

CI, confidence interval; mFAS, modified full analysis set.

#### B. Subgroups: Protocol-defined risk factors

| Risk Factor (Sub) Group | REGEN-COV Events/N (%) | Placebo Events/N (%) | Rel. Risk Reduction (95% CI) | P |
|---|---|---|---|---|
| **>=1 protocol-defined risk factor for severe COVID-19** | | | | |
| 1200 mg vs Placebo | 7/736 ( 1.0) | 24/748 ( 3.2) | 70% (32%, 87%) | 0.0024 |
| 2400 mg vs Placebo | 18/1355 ( 1.3) | 62/1341 ( 4.6) | 71% (52%, 83%) | <0.0001 |
| **Age >= 50 years** | | | | |
| 1200 mg vs Placebo | 7/357 ( 2.0) | 19/356 ( 5.3) | 63% (14%, 84%) | 0.0166 |
| 2400 mg vs Placebo | 15/715 ( 2.1) | 52/678 ( 7.7) | 73% (52%, 84%) | <0.0001 |
| **Obesity (BMI >= 30 kg/m^2)** | | | | |
| 1200 mg vs Placebo | 2/410 ( 0.5) | 15/427 ( 3.5) | 86% (40%, 97%) | 0.0022 |
| 2400 mg vs Placebo | 13/787 ( 1.7) | 38/772 ( 4.9) | 66% (38%, 82%) | 0.0003 |
| **Cardiovascular disease, including hypertension** | | | | |
| 1200 mg vs Placebo | 5/282 ( 1.8) | 15/266 ( 5.6) | 69% (15%, 88%) | 0.0211 |
| 2400 mg vs Placebo | 7/520 ( 1.3) | 40/473 ( 8.5) | 84% (65%, 93%) | <0.0001 |
| **Chronic metabolic disease, including diabetes** | | | | |
| 1200 mg vs Placebo | 2/94 ( 2.1) | 6/100 ( 6.0) | 64% (NA, 93%) | 0.2807 |
| 2400 mg vs Placebo | 8/202 ( 4.0) | 17/210 ( 8.1) | 51% (NA, 78%) | 0.0842 |
| **Chronic liver disease** | | | | |
| 1200 mg vs Placebo | 0/3 ( 0.0) | 0/4 ( 0.0) | NA (NA, NA) | NA |
| 2400 mg vs Placebo | 0/14 ( 0.0) | 1/8 (12.5) | 100% (NA, NA) | 0.3636 |
| **Chronic kidney disease, including those on dialysis** | | | | |
| 1200 mg vs Placebo | 1/8 (12.5) | 1/4 (25.0) | 50% (NA, 96%) | 1 |
| 2400 mg vs Placebo | 2/19 (10.5) | 4/9 (44.4) | 76% (NA, 95%) | 0.0638 |
| **Chronic lung disease, including asthma** | | | | |
| 1200 mg vs Placebo | 1/139 ( 0.7) | 6/139 ( 4.3) | 83% (NA, 98%) | 0.1203 |
| 2400 mg vs Placebo | 5/216 ( 2.3) | 14/219 ( 6.4) | 64% (1%, 87%) | 0.058 |
| **Immunosuppressed** | | | | |
| 1200 mg vs Placebo | 0/24 ( 0.0) | 1/10 (10.0) | 100% (NA, NA) | 0.2941 |
| 2400 mg vs Placebo | 2/46 ( 4.3) | 2/34 ( 5.9) | 26% (NA, 89%) | 1 |

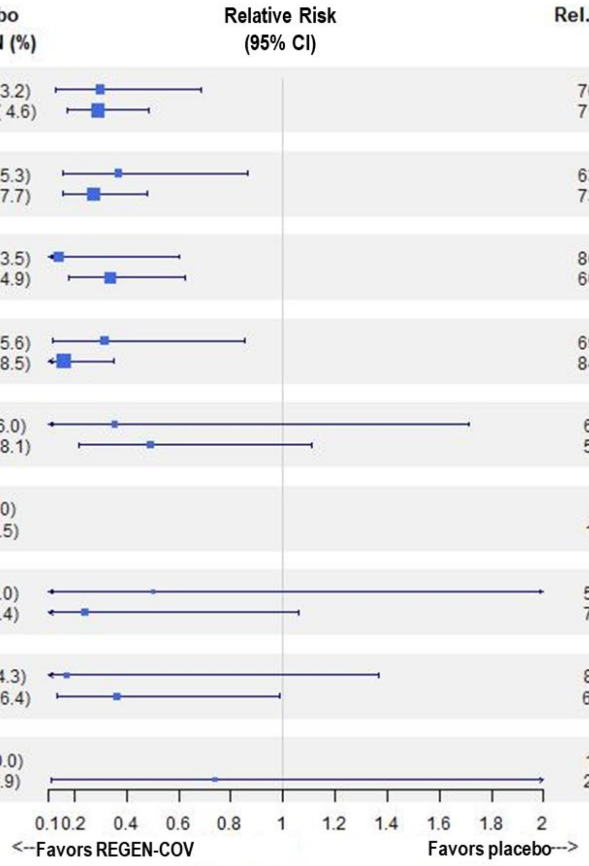

BMI, body mass index; CI, confidence interval.

#### C. Subgroups: Other risk factor combinations

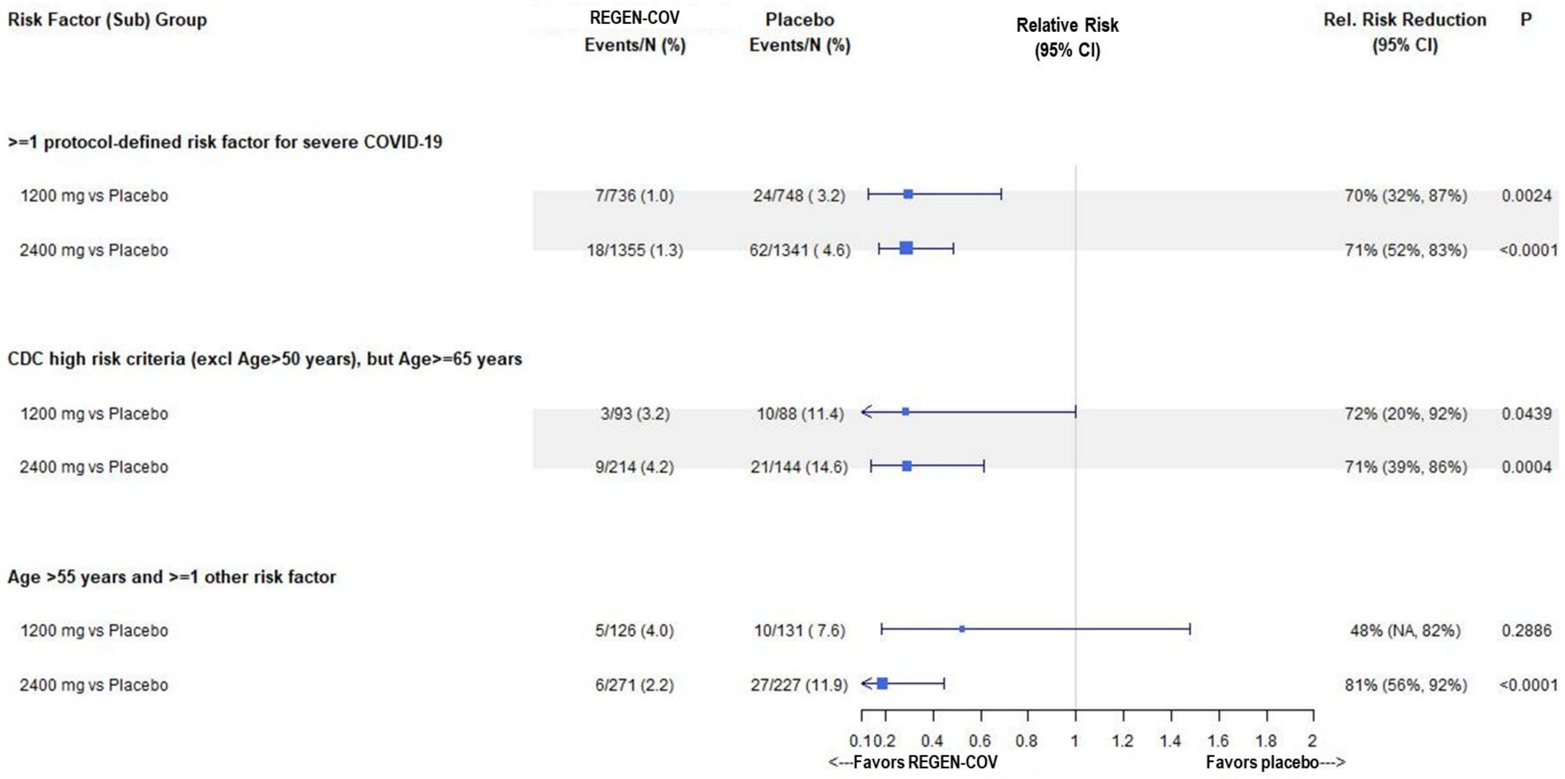

CDC, Centers for Disease Control and Prevention; CI, confidence interval.

#### Figure S4. COVID-19-related Hospitalization or All-Cause Death – Day 4 through Day 29

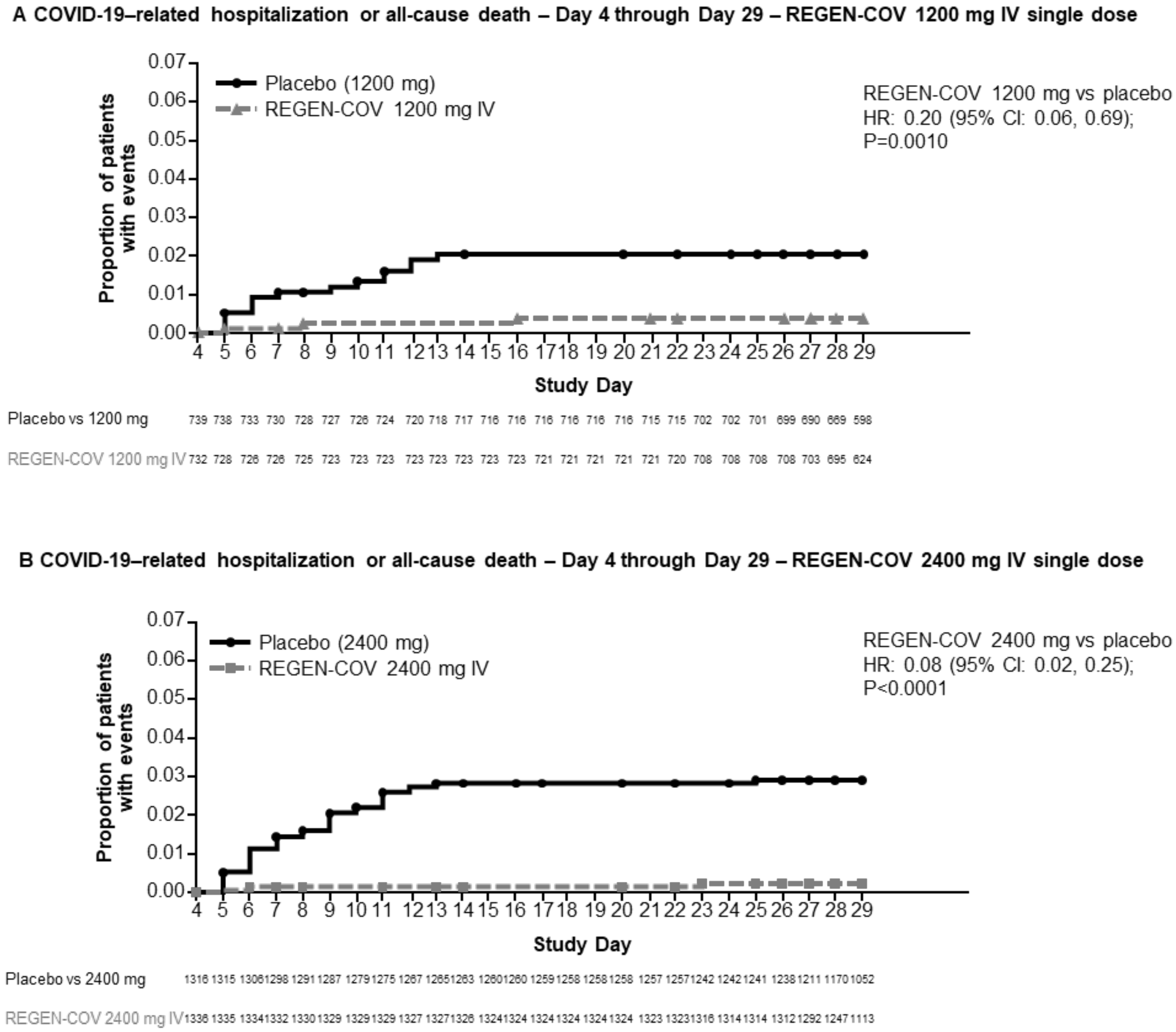

CI, confidence interval; HR, hazard ratio; IV, intravenous.

### Figure S5. Forest Plot: Time to Symptoms Resolution

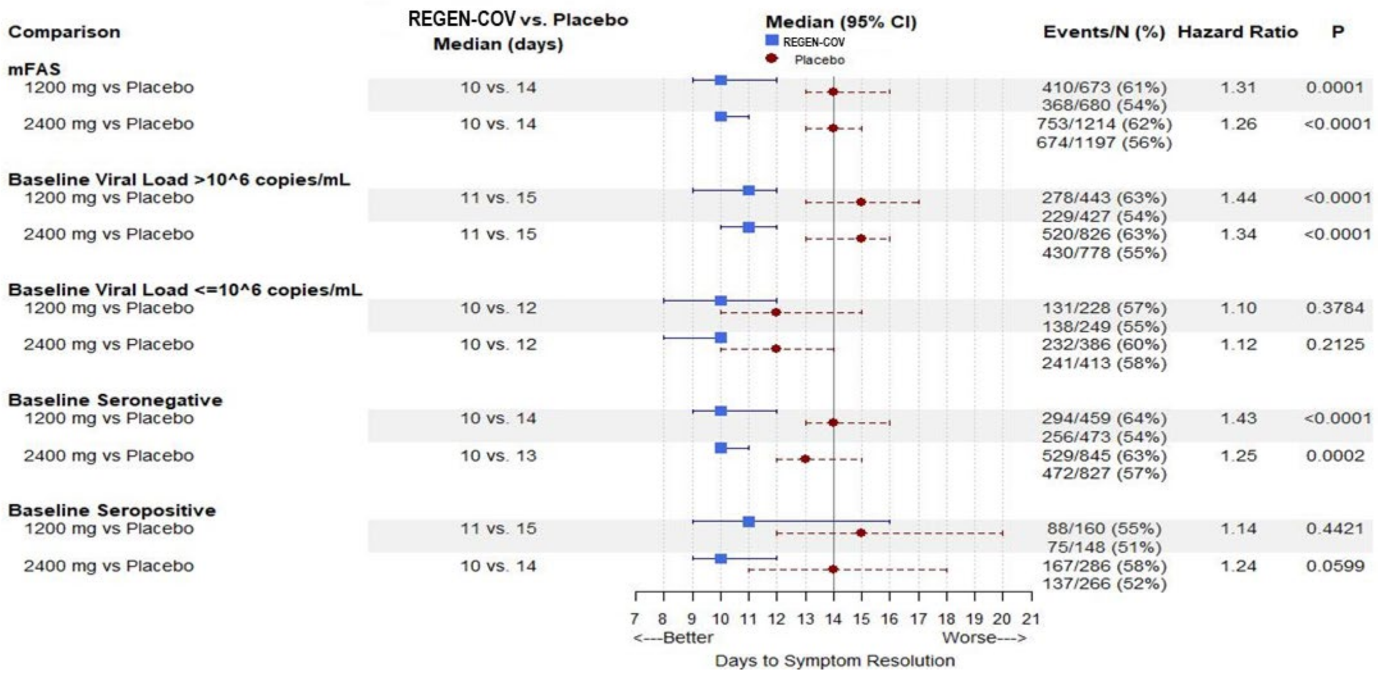

CI, confidence interval; mFAS, modified full analysis set.

#### Figure S6. Virologic Efficacy – Amended Phase 3 Portion

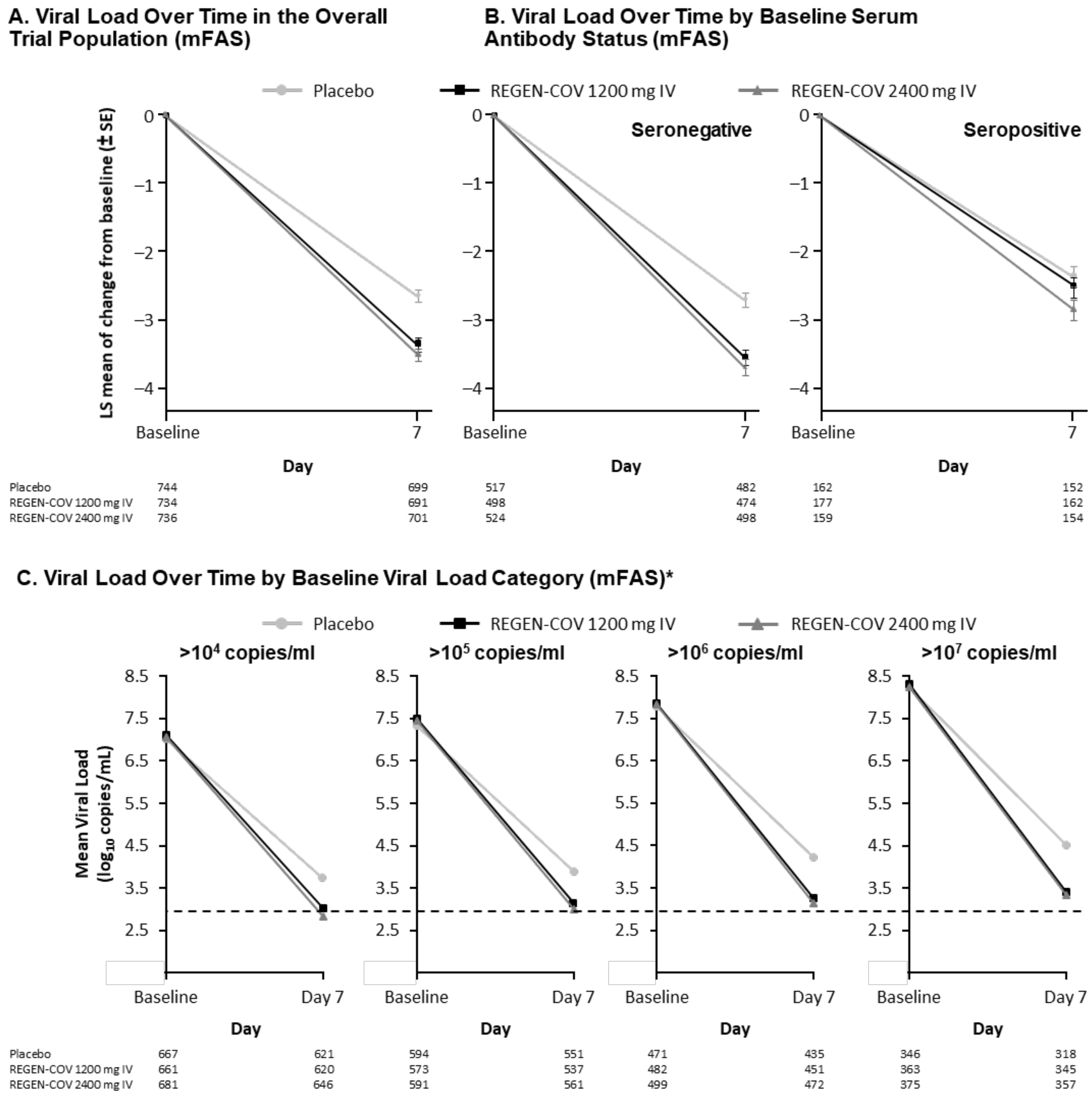

* The lower limit of detection (dashed line) is 714 copies per milliliter (2.85 $\log_{10}$ copies per milliliter).

IV, intravenous(ly); mFAS, modified full analysis set; SE, standard error.

.

#### Figure S7. Forest Plot: Virologic Efficacy – Amended Phase 3 Portion

| | REGEN-COV LSMean (SE) | Placebo LSMean (SE) | Difference (95% CI) | P Value |
|---|---|---|---|---|
| **mFAS** | | | | |
| 1200 mg vs Placebo (n=1484) | -3.35 (0.09) | -2.64 (0.09) | -0.71 (-0.90, -0.53) | <0.0001 |
| 2400 mg vs Placebo (n=2696) | -3.32 (0.09) | -2.47 (0.09) | -0.86 (-1.00, -0.72) | <0.0001 |
| **Baseline Viral Load >10^6 copies/mL** | | | | |
| 1200 mg vs Placebo (n=953) | -4.09 (0.12) | -3.07 (0.12) | -1.01 (-1.24, -0.79) | <0.0001 |
| 2400 mg vs Placebo (n=1800) | -3.91 (0.12) | -2.87 (0.12) | -1.04 (-1.20, -0.87) | <0.0001 |
| **Baseline Seronegative** | | | | |
| 1200 mg vs Placebo (n=963) | -3.56 (0.11) | -2.70 (0.11) | -0.86 (-1.09, -0.64) | <0.0001 |
| 2400 mg vs Placebo (n=1870) | -3.58 (0.11) | -2.55 (0.11) | -1.04 (-1.20, -0.87) | <0.0001 |
| **Baseline Seropositive** | | | | |
| 1200 mg vs Placebo (n=341) | -2.53 (0.15) | -2.36 (0.16) | -0.16 (-0.53, 0.20) | 0.3799 |
| 2400 mg vs Placebo (n=620) | -2.36 (0.18) | -1.94 (0.19) | -0.43 (-0.70, -0.15) | 0.0027 |

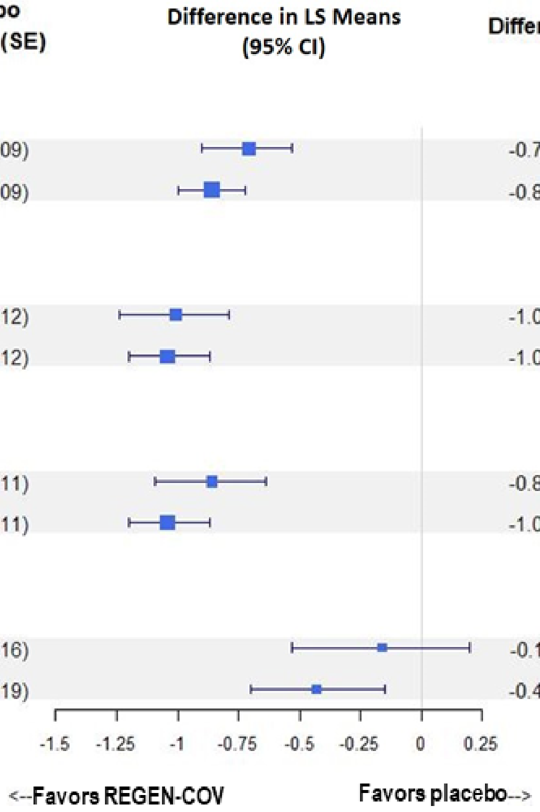

CI, confidence interval; LS, least squares; mFAS, modified full analysis set; SE, standard error.

### Figure S8. Virologic Efficacy – Original Phase 3 Portion

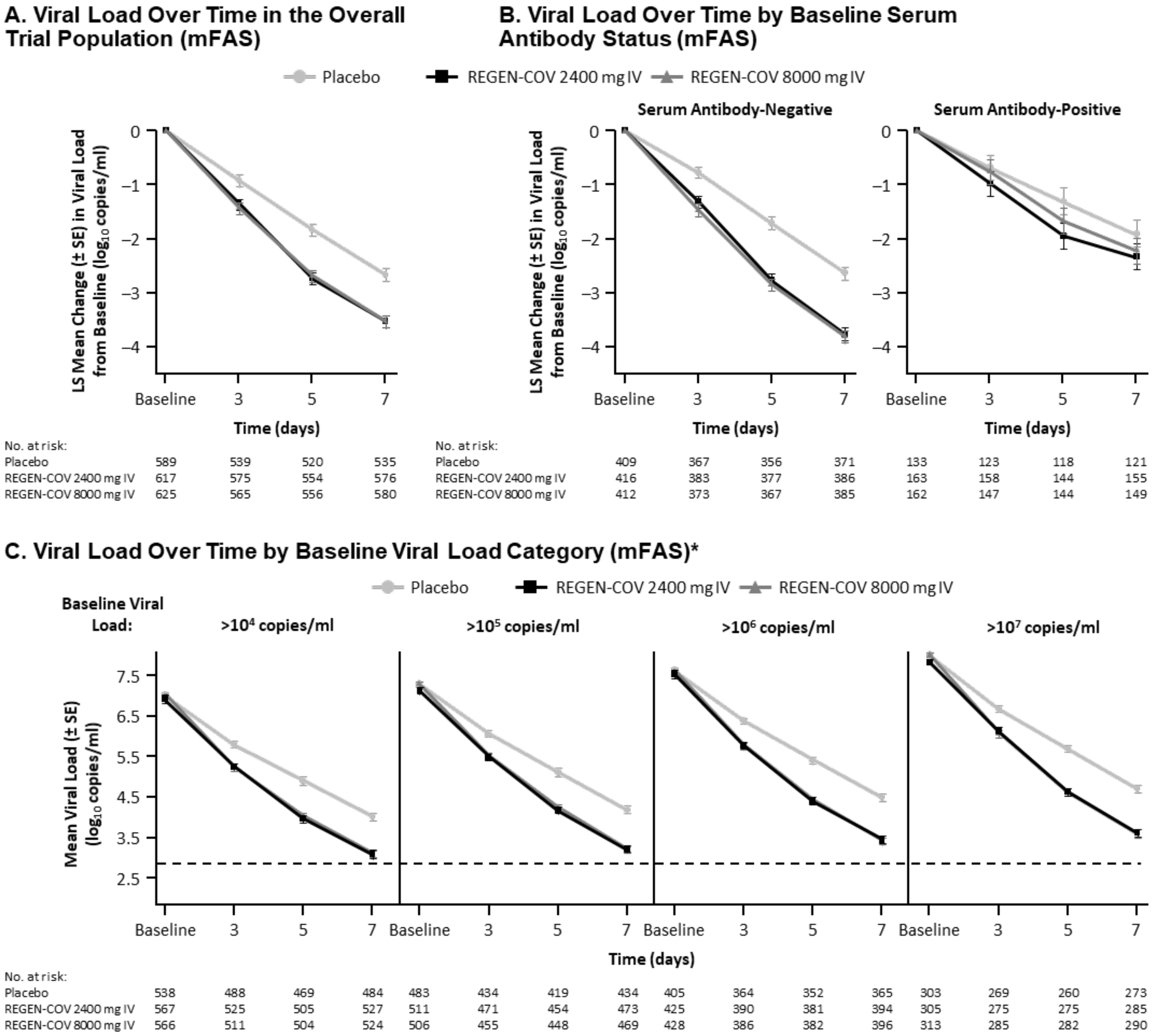

* The lower limit of detection (dashed line) is 714 copies per milliliter (2.85 log10 copies per milliliter).

mFAS, modified full analysis set; IV, intravenous, SE, standard error.

#### Supplementary Tables

##### Table S1. Phase 3 Primary Analysis Hierarchical Testing Order

The analysis of the primary endpoint (proportion of patients with ≥1 COVID-19-related hospitalization or all-cause death through day 29) and key secondary endpoint (time to symptom resolution) will be conducted at the overall α=0.05. The endpoints will be tested hierarchically in the following order, adjusting for interim analysis:

| Hierarchy Number | Description |
| --- | --- |
| 1 | Proportion of patients with ≥1 COVID-19-related hospitalization or all-cause death through day 29 in the mFAS for REGEN-COV 2400 mg group versus placebo |
| 2 | Proportion of patients with ≥1 COVID-19-related hospitalization or all-cause death through day 29 in the mFAS for REGEN-COV 1200 mg group versus placebo |
| 3 | Proportion of patients with ≥1 COVID-19-related hospitalization or all-cause death through day 29 in the mFAS patients with baseline viral load >$10^6$ copies/mL for REGEN-COV 2400 mg group versus placebo |
| 4 | Proportion of patients with ≥1 COVID-19-related hospitalization or all-cause death through day 29 in the mFAS patients who are seronegative at baseline for REGEN-COV 2400 mg group versus placebo |
| 5 | Proportion of patients with ≥1 COVID-19-related hospitalization or all-cause death through day 29 in the mFAS patients with baseline viral load >$10^6$ copies/mL for REGEN-COV 1200 mg group versus placebo |
| 6 | Proportion of patients with ≥1 COVID-19-related hospitalization or all-cause death through day 29 in the mFAS patients who are seronegative at baseline for REGEN-COV 1200 mg group versus placebo |
| 7 | Proportion of patients with ≥1 COVID-19-related hospitalization or all-cause death from day 4 through day 29 in the mFAS for REGEN-COV 2400 mg group versus placebo |
| 8 | Proportion of patients with ≥1 COVID-19-related hospitalization or all-cause death from day 4 through day 29 in the mFAS for REGEN-COV 1200 mg group versus placebo |
| 9 | Time to COVID-19 symptoms resolution in the mFAS for REGEN-COV 2400 mg group versus placebo |
| 10 | Time to COVID-19 symptoms resolution in the mFAS for REGEN-COV 1200 mg group versus placebo |

mFAS, modified full analysis set.

#### Table S2. Protocol-Defined Risk Factors for Severe Covid-19 (mFAS)

| **Protocol-defined risk factor** — no. (%) | **Placebo (n=1341)** | **REGEN-COV 1200 mg (n=736)** | **REGEN-COV 2400 mg (n=1355)** | **REGEN-COV 8000 mg (n=625)** | **Total (n=4057)** |
|---|---|---|---|---|---|
| Age ≥50 years | 678 (50.6) | 357 (48.5) | 715 (52.8) | 351 (56.2) | 2101 (51.8) |
| Obesity (BMI ≥30 kg/m$^2$) | 772 (57.6) | 410 (55.7) | 787 (58.1) | 384 (61.4) | 2353 (58.0) |
| Cardiovascular disease, including hypertension | 473 (35.3) | 282 (38.3) | 520 (38.4) | 196 (31.4) | 1471 (36.3) |
| Chronic lung disease, including asthma | 219 (16.3) | 139 (18.9) | 216 (15.9) | 92 (14.7) | 666 (16.4) |
| Type 1 or 2 diabetes mellitus | 210 (15.7) | 94 (12.8) | 202 (14.9) | 97 (15.5) | 603 (14.9) |
| Chronic kidney disease, including those on dialysis | 9 (0.7) | 8 (1.1) | 19 (1.4) | 9 (1.4) | 45 (1.1) |
| Chronic liver disease | 8 (0.6) | 3 (0.4) | 14 (1.0) | 11 (1.8) | 36 (0.9) |
| Immunocompromised* | 34 (2.5) | 24 (3.3) | 46 (3.4) | 16 (2.6) | 120 (3.0) |

BMI, body mass index; mFAS, modified full analysis set.

* The most common immunosuppressive conditions were rheumatoid arthritis, HIV/AIDS, and systemic lupus erythematosus; the most common immunosuppressive medications were hydroxychloroquine, antimetabolites, and TNF inhibitors.

#### Table S3. Demographic and Baseline Medical Characteristics (mFAS) – 8000 mg REGEN-COV

| Characteristic* | REGEN-COV 8000 mg (n=625) | Placebo (8000 mg) (concurrent) (n=593) |
|---|---|---|
| **Demographics** | | |
| Median age (IQR) — year | 51.0 (40.0–59.0) | 50.0 (39.0–58.0) |
| Baseline age category — no. (%) | | |
| Age ≥50 years | 351 (56.2) | 322 (54.3) |
| Age ≥65 years | 97 (15.5) | 56 (9.4) |
| Male sex — no. (%) | 324 (51.8) | 281 (47.4) |
| Hispanic or Latino ethnic group — no. (%)† | 177 (28.3) | 176 (29.7) |
| Race — no. (%)† | | |
| White | 534 (85.4) | 525 (88.5) |
| Black or African American | 33 (5.3) | 28 (4.7) |
| Asian | 26 (4.2) | 20 (3.4) |
| American Indian or Alaska Native | 3 (0.5) | 3 (0.5) |
| Unknown | 15 (2.4) | 6 (1.0) |
| Not reported | 14 (2.2) | 11 (1.9) |
| Median weight (IQR) — kg | 89.85 (76.20–106.60) | 88.50 (75.00–104.00) |

| | | |
|---|---|---|
| Body-mass index‡ | 31.90±7.229 | 31.35±6.846 |
| Obesity — no. (%)§ | 384 (61.4) | 345 (58.2) |
| At least one risk factor for severe Covid-19 — no. (%)¶ | 625 (100) | 593 (100) |
| **Medical/Clinical Characteristics** | | |
| Positive baseline qualitative RT-PCR — no. (%) | 625 (100) | 589 (99.3) |
| Baseline viral load in nasopharyngeal swab (raw values) | | |
| No. of patients | 625 | 589 |
| Mean viral load — (10^6) copies/ml | 170.03 ± 555.7 | 194.01 ± 628.9 |
| Median viral load (range) — (10^6) copies/ml | 10.10 (0–6090) | 11.20 (0–6780) |
| Baseline viral load in nasopharyngeal swab ($log_{10}$ scale) | | |
| No. of patients | 625 | 589 |
| Mean viral load — $log_{10}$ copies/ml | 6.64 ± 1.671 | 6.70 ± 1.664 |
| Median viral load (range) — $log_{10}$ copies/ml | 7.00 (2.6–9.8) | 7.05 (2.6–9.8) |
| Baseline serum C-reactive protein level | | |
| No. of patients | 544 | 519 |
| Mean level — mg/l | 14.206 ± 28.0260 | 12.794 ± 23.9368 |
| Median level (range) — mg/l | 5.065 (0.18–228.07) | 5.020 (0.10–242.73) |

| | | |
|---|---|---|
| Baseline serum antibody status — no. (%) | | |
| Negative | 412 (65.9) | 411 (69.3) |
| Positive | 162 (25.9) | 133 (22.4) |
| Other | 51 (8.2) | 49 (8.3) |
| Median time from symptom onset to randomization (IQR) — days | 3.0 (2–5) | 3.0 (2–5) |

* Plus–minus values are means ±SD. Percentages may not total 100 because of rounding. IQR denotes interquartile range, RT-PCR reverse-transcriptase polymerase chain reaction, and SD standard deviation.
† Race and ethnic group were reported by the patients.
‡ The body-mass index is the weight in kilograms divided by the square of the height in meters.
§ Obesity is defined as a body-mass index of greater than or equal to 30.
¶ Risk factors for hospitalization include an age of more than 50 years, obesity, cardiovascular disease (including hypertension), chronic lung disease (including asthma), chronic metabolic disease (including diabetes), chronic kidney disease (including receipt of dialysis), chronic liver disease, and immunocompromised (immunosuppression or receipt of immunosuppressants).

#### Table S4. Proportion of Patients in the Placebo Arms with ≥1 Covid-19-related Hospitalization or All-cause Death by Baseline Viral Load Category and Baseline Serum Antibody Status

| End Point | Placebo (concurrent with REGEN-COV 2400 mg) | Placebo (concurrent with REGEN-COV 1200 mg) |
|---|---|---|
| **Proportion of patients with ≥1 Covid-19-related hospitalization or all-cause death** | | |
| **Baseline viral load category: high viral load (>$10^6$ copies/mL)** | | |
| No. of patients | 876 | 471 |
| Patients with event within 29 days — no. (%) | 55 (6.3) | 20 (4.2) |
| **Baseline viral load category: low viral load (≤$10^6$ copies/mL)** | | |
| No. of patients | 457 | 273 |
| Patients with event within 29 days — no. (%) | 6 (1.3) | 4 (1.5) |
| **Proportion of patients with ≥1 Covid-19-related hospitalization or all-cause death** | | |
| **Baseline serum antibody status: negative** | | |
| No. of patients | 930 | 519 |
| Patients with event within 29 days — no. (%) | 49 (5.3) | 18 (3.5) |
| **Baseline serum antibody status: positive** | | |
| No. of patients | 297 | 164 |
| Patients with event within 29 days — no. (%) | 12 (4.0) | 6 (3.7) |
| **Baseline serum antibody status: other** | | |
| No. of patients | 114 | 65 |
| Patients with event within 29 days — no. (%) | 1 (0.9) | 0 |

#### Table S5. Viral Load in the Placebo Arm by With and Without Hospitalization or Death and by Baseline Serum Antibody Status

| Baseline Serum Antibody Status: | Covid-19-related Hospitalization or Death | Baseline SARS-CoV-2 Viral Load ($log_{10}$ copies/ml) (Mean ± SD) | Day 7 SARS-CoV-2 Viral Load ($log_{10}$ copies/ml) (Mean ± SD) |
|---|---|---|---|
| **All** | | | |
| n=1272 | No | 6.62 ± 1.77 | 3.60 ± 2.13 |
| n=61 | Yes | 7.54 ± 1.18 | 5.36 ± 1.35 |
| **Negative** | | | |
| n=877 | No | 7.16 ± 1.49 | 4.03 ± 2.01 |
| n=48 | Yes | 7.61 ± 1.09 | 5.41 ± 1.36 |
| **Positive** | | | |
| n=283 | No | 5.04 ± 1.62 | 2.46 ± 2.04 |
| n=12 | Yes | 7.06 ± 1.35 | 5.08 ± 1.39 |

SD, standard deviation.

#### Table S6. Proportion of Patients with ≥1 Covid-19-related Hospitalization or All-cause Death

| End Point | REGEN-COV 2400 mg (n=1355) | Placebo (2400 mg) (concurrent) (n=1341) | REGEN-COV 1200 mg (n=736) | Placebo (1200 mg) (concurrent) (n=748) |
|---|---|---|---|---|
| **Proportion of patients with ≥1 Covid-19-related hospitalization or all-cause death** | | | | |
| Patients with event within 29 days — no. (%) | 18 (1.3) | 62 (4.6) | 7 (1.0) | 24 (3.2) |
| Relative risk reduction vs placebo — percentage points | 71.3 | | 70.4 | |
| 95% CI† | 51.7, 82.9 | | 31.6, 87.1 | |
| **Proportion of patients with hospitalization** | | | | |
| Patients with event within 29 days — no. (%) | 17 (1.3) | 59 (4.4) | 6 (0.8) | 23 (3.1) |
| Relative risk reduction vs placebo — percentage points | 71.5 | | 73.5 | |
| 95% CI† | 51.3, 83.3 | | 35.3, 89.1 | |
| **Proportion of patients with all-cause death** | | | | |
| Patients with event within 29 days — no. (%) | 1 (<0.1) | 3 (0.2) | 1 (0.1) | 1 (0.1) |
| Relative risk reduction vs placebo — percentage points | 67.0 | | -1.6 | |
| 95% CI† | -216.7, 96.6 | | -1522, 93.6 | |

CI, confidence interval.

† 95% CI used the Farrington-Manning method.

#### Table S7. Proportion of Patients with ≥1 Covid-19-related MAV or All-cause Death

| End Point* | REGEN-COV 2400 mg (n=1355) | Placebo (2400 mg) (concurrent) (n=1341) | REGEN-COV 1200 mg (n=736) | Placebo (1200 mg) (concurrent) (n=748) |
|---|---|---|---|---|
| **Proportion of patients with ≥1 Covid-19-related MAV or all-cause death** | | | | |
| Patients with event within 29 days — no. (%) | 43 (3.2) | 109 (8.1) | 20 (2.7%) | 51 (6.8) |
| Relative risk reduction vs placebo — percentage points | 61.0 | | 60.1 | |
| 95% CI† | 44.9, 72.3 | | 33.8, 76.0 | |
| **Proportion of patients with hospitalization** | | | | |
| Patients with event within 29 days — no. (%) | 17 (1.3) | 59 (4.4) | 6 (0.8) | 23 (3.1) |
| Relative risk reduction vs placebo — percentage points | 71.5 | | 73.5 | |
| 95% CI† | 51.3, 83.3 | | 35.3, 89.1 | |
| **Proportion of patients with emergency room visit** | | | | |
| Patients with event within 29 days — no. (%) | 9 (0.7) | 16 (1.2) | 2 (0.3) | 10 (1.3) |
| Relative risk reduction vs placebo — percentage points | 44.3 | | 79.7 | |
| 95% CI† | -25.5, 75.3 | | 7.5, 95.5 | |
| **Proportion of patients with urgent care visit** | | | | |
| Patients with event within 29 days — no. (%) | 3 (0.2) | 7 (0.5) | 1 (0.1) | 5 (0.7) |
| Relative risk reduction vs placebo — percentage points | 57.6 | | 79.7 | |
| 95% CI† | -63.7, 89.0 | | -73.6, 97.6 | |

| | | | | |
|---|---|---|---|---|
| **Proportion of patients with physician office/telemedicine visit** | | | | |
| Patients with event within 29 days — no. (%) | 13 (1.0) | 24 (1.8) | 10 (1.4) | 12 (1.6) |
| Relative risk reduction vs placebo — percentage points | 46.4 | | 15.3 | |
| 95% CI† | -4.8, 72.6 | | -94.8, 63.2 | |
| **Proportion of patients with all-cause death** | | | | |
| Patients with event within 29 days — no. (%) | 1 (<0.1) | 3 (0.2) | 1 (0.1) | 1 (0.1) |
| Relative risk reduction vs placebo — percentage points | 67.0 | | -1.6 | |
| 95% CI† | -216.7, 96.6 | | -1522, 93.6 | |

CI, confidence interval; MAV, medically-attended visit.

* A patient with multiple types of events was only counted to the worst level event, following this decreasing order: all-cause death, hospitalization, ER, urgent care, physician office visit/telemedicine.
† 95% CI used the Farrington-Manning method.

#### Table S8. Hospitalization Outcomes: Length of Stay, ICU, and Mechanical Ventilation

| End Point | REGEN-COV 2400 mg (n=1355) | Placebo (2400 mg) (concurrent) (n=1341) | REGEN-COV 1200 mg (n=736) | Placebo (1200 mg) (concurrent) (n=748) |
|---|---|---|---|---|
| **Days of hospitalization due to COVID-19 per patient** | | | | |
| No. of patients | 18 | 62 | 7 | 24 |
| Mean (SD) | 8.6 ± 7.07 | 10.0 ± 7.16 | 7.0 ± 8.04 | 8.4 ± 6.74 |
| Median (IQR) | 6.0 (3.0–11.0) | 7.0 (5.0–13.0) | 4.0 (3.0–6.0) | 5.5 (4.0–10.5) |
| **Proportion of patients admitted to an ICU** | | | | |
| Patients with event within 29 days — no. (%) | 6 (0.4) | 18 (1.3) | 3 (0.4) | 7 (0.9) |
| Relative risk reduction vs placebo — percentage points | 67.0 | | 56.4 | |
| 95% CI* | 17.2, 86.9 | | -67.8, 88.7 | |
| **Proportion of patients requiring mechanical ventilation** | | | | |
| Patients with event within 29 days — no. (%) | 1 (<0.1) | 6 (0.4) | 1 (0.1) | 2 (0.3) |
| Relative risk reduction vs placebo — percentage points | 83.5 | | 49.2 | |
| 95% CI* | -36.8, 98.0 | | -459.2, 95.4 | |

CI, confidence interval; ICU, intensive care unit, IQR, interquartile range, SD, standard deviation.

* 95% CI used the Farrington-Manning method.

#### Table S9. Proportion of Patients with ≥1 Covid-19-related Hospitalization, Emergency Room Visits, or All-cause Death

| End Point | REGEN-COV 2400 mg (n=1355) | Placebo (2400 mg) (concurrent) (n=1341) | REGEN-COV 1200 mg (n=736) | Placebo (1200 mg) (concurrent) (n=748) |
|---|---|---|---|---|
| **Proportion of patients with events** | | | | |
| Patients with event within 29 days — no. (%) | 27 (2.0) | 78 (5.8) | 9 (1.2) | 34 (4.5) |
| Relative risk reduction vs placebo — percentage points | 65.7 | | 73.1 | |
| 95% CI* | 47.3, 77.7 | | 44.3, 87.0 | |

CI, confidence interval.

* 95% CI used the Farrington-Manning method.

#### Table S10. Proportion of Patients with ≥1 Covid-19-related Hospitalization or All-cause Death – 8000 mg REGEN-COV

| End Point | REGEN-COV 8000 mg (n=625) | Placebo (8000 mg) (concurrent) (n=593) |
|---|---|---|
| **Proportion of patients with ≥1 COVID-19-related hospitalization or all-cause death** | | |
| Patients with event within 29 days — no. (%) | 13 (2.1) | 38 (6.4) |
| Relative risk reduction vs placebo — percentage points | 67.5 | |
| 95% CI† | 39.7, 82.5 | |
| **Proportion of patients with hospitalization** | | |
| Patients with event within 29 days — no. (%) | 13 (2.1) | 36 (6.1) |
| Relative risk reduction vs placebo — percentage points | 65.7 | |
| 95% CI† | 36.0, 81.6 | |
| **Proportion of patients with all-cause death** | | |
| Patients with event within 29 days — no. (%) | 0 | 2 (0.3) |
| Relative risk reduction vs placebo — percentage points | 100.0 | |
| 95% CI† | n/a | |

CI, confidence interval.

† 95% CI used the Farrington-Manning method.

#### Table S11. Proportion of Patients with ≥1 Covid-19-related MAV or All-cause Death – 8000 mg REGEN-COV

| End Point* | REGEN-COV 8000 mg (n=625) | Placebo (8000 mg) (concurrent) (n=593) |
|---|---|---|
| **Proportion of patients with ≥1 Covid-19-related MAV or all-cause death** | | |
| Patients with event within 29 days — no. (%) | 26 (4.2) | 58 (9.8) |
| Relative risk reduction vs placebo — percentage points | 57.5 | |
| 95% CI† | 33.4, 72.8 | |
| **Proportion of patients with hospitalization** | | |
| Patients with event within 29 days — no. (%) | 13 (2.1) | 36 (6.1) |
| Relative risk reduction vs placebo — percentage points | 65.7 | |
| 95% CI† | 36.0, 81.6 | |
| **Proportion of patients with emergency room visit** | | |
| Patients with event within 29 days — no. (%) | 3 (0.5) | 6 (1.0) |
| Relative risk reduction vs placebo — percentage points | 52.6 | |
| 95% CI† | -88.8, 88.1 | |
| **Proportion of patients with urgent care visit** | | |
| Patients with event within 29 days — no. (%) | 2 (0.3) | 2 (0.3) |
| Relative risk reduction vs placebo — percentage points | 5.1 | |

| | | |
|---|---|---|
| 95% CI† | -571.4, 86.6 | |
| **Proportion of patients with physician office/telemedicine visit** | | |
| Patients with event within 29 days — no. (%) | 8 (1.3) | 12 (2.0) |
| Relative risk reduction vs placebo — percentage points | 36.7 | |
| 95% CI† | -53.6, 74.0 | |
| **Proportion of patients with all-cause death** | | |
| Patients with event within 29 days — no. (%) | 0 | 2 (0.3) |
| Relative risk reduction vs placebo — percentage points | 100.0 | |
| 95% CI† | n/a | |

CI, confidence interval; MAV, medically-attended visit.

* A patient with multiple types of events was only counted to the worst level event, following this decreasing order: all-cause death, hospitalization, ER, urgent care, physician office visit/telemedicine.
† 95% CI used the Farrington-Manning method.

#### Table S12. Treatment-Emergent Adverse Events Leading to Death

| **System Organ Class**<br>Preferred Term | **Placebo (n=1476)** | **REGEN-COV 1200 mg IV (n=827)** | **REGEN-COV 2400 mg IV (n=1512)** | **REGEN-COV 8000 mg IV (n=689)** | **Total (n=4504)** |
|---|---|---|---|---|---|
| *no. of patients (percent)* | | | | | |
| **Number of patients with at least one treatment-emergent adverse event leading to death** | | | | | |
| TEAE leading to death | 5 (0.3) | 1 (0.1) | 1 (<0.1) | 0 | 7 (0.2) |
| **Treatment-emergent adverse event leading to death by system organ class and preferred term** | | | | | |
| **Respiratory, thoracic and mediastinal disorders** | | | | | |
| Dyspnoea | 1 (<0.1) | 0 | 1 (<0.1) | 0 | 2 (<0.1) |
| Acute respiratory distress syndrome | 1 (<0.1) | 0 | 0 | 0 | 1 (<0.1) |
| Hypoxia | 0 | 1 (0.1) | 0 | 0 | 1 (<0.1) |
| Respiratory failure | 1 (<0.1) | 0 | 0 | 0 | 1 (<0.1) |
| **Infections and infestations** | | | | | |
| COVID-19 | 1 (<0.1) | 0 | 0 | 0 | 1 (<0.1) |
| COVID-19 pneumonia | 1 (<0.1) | 0 | 0 | 0 | 1 (<0.1) |
| Pneumonia | 1 (<0.1) | 0 | 0 | 0 | 1 (<0.1) |
| **Neoplasms benign, malignant and unspecified (including cysts and polyps)** | | | | | |
| Tumor obstruction | 1 (<0.1) | 0 | 0 | 0 | 1 (<0.1) |

IV, intravenous(ly).

#### Table S13. Treatment-Emergent Serious Adverse Events and Adverse Events of Special Interest

| **System Organ Class**<br>Preferred Term* | **Placebo (n=1843)** | **REGEN-COV 1200 mg IV (n=827)** | **REGEN-COV 2400 mg IV (n=1849)** | **REGEN-COV 8000 mg IV (n=1012)** | **Total (n=5531)** |
|---|---|---|---|---|---|
| | *no. of patients (percent)*† | | | | |
| **Serious treatment-emergent adverse events by system organ class and preferred term** | | | | | |
| **Infections and infestations** | | | | | |
| COVID-19 | 18 (1.0%) | 1 (0.1%) | 5 (0.3%) | 5 (0.5%) | 29 (0.5%) |
| COVID-19 pneumonia | 14 (0.8%) | 2 (0.2%) | 4 (0.2%) | 5 (0.5%) | 25 (0.5%) |
| Pneumonia | 17 (0.9%) | 2 (0.2%) | 3 (0.2%) | 1 (<0.1%) | 23 (0.4%) |
| **Respiratory, thoracic and mediastinal disorders** | | | | | |
| Dyspnoea | 7 (0.4%) | 0 | 1 (<0.1%) | 1 (<0.1%) | 9 (0.2%) |
| Hypoxia | 6 (0.3%) | 1 (0.1%) | 1 (<0.1%) | 1 (<0.1%) | 9 (0.2%) |
| Acute respiratory failure | 3 (0.2%) | 0 | 2 (0.1%) | 1 (<0.1%) | 6 (0.1%) |
| Respiratory distress | 2 (0.1%) | 0 | 0 | 0 | 2 (<0.1%) |
| **Metabolism and nutrition disorders** | | | | | |
| Dehydration | 2 (0.1%) | 0 | 0 | 0 | 2 (<0.1%) |
| Hyponatraemia | 2 (0.1%) | 0 | 0 | 0 | 2 (<0.1%) |
| **Adverse events of special interest by preferred term** | | | | | |
| COVID-19 | 11 (0.6%) | 2 (0.2%) | 4 (0.2%) | 2 (0.2%) | 19 (0.3%) |
| Dyspnoea | 9 (0.5%) | 3 (0.4%) | 3 (0.2%) | 1 (<0.1%) | 16 (0.3%) |
| Cough | 2 (0.1%) | 3 (0.4%) | 2 (0.1%) | 1 (<0.1%) | 8 (0.1%) |
| Pneumonia | 6 (0.3%) | 2 (0.2%) | 0 | 0 | 8 (0.1%) |
| COVID-19 pneumonia | 4 (0.2%) | 2 (0.2%) | 0 | 1 (<0.1%) | 7 (0.1%) |
| Headache | 2 (0.1%) | 2 (0.2%) | 1 (<0.1%) | 1 (<0.1%) | 6 (0.1%) |
| Dizziness | 1 (<0.1%) | 2 (0.2%) | 1 (<0.1%) | 0 | 4 (<0.1%) |
| Nausea | 0 | 2 (0.2%) | 0 | 1 (<0.1%) | 3 (<0.1%) |
| Pulmonary congestion | 1 (<0.1%) | 0 | 0 | 2 (0.2%) | 3 (<0.1%) |
| Nasal congestion | 2 (0.1%) | 0 | 0 | 0 | 2 (<0.1%) |

* Term included if ≥2 patients in any of the individual dose groups.

† A patient who reported 2 or more adverse events with different preferred terms within the same system organ class is counted only once in that system organ class. A patient who reported 2 or more adverse events with the same preferred term is counted only once for that term. If a patient had more than one occurrence in the same event category, only the most related was counted.

IV, intravenous(ly).

#### Table S14. Mean (SD) [N] Pharmacokinetic Parameters of REGN10933 and REGN10987 in Serum

| Pharmacokinetic Parameter | REGN10933 (casirivimab)* | | | REGN10987 (imdevimab)* | | |
|---|---|---|---|---|---|---|
| | 600 mg | 1200 mg | 4000 mg | 600 mg | 1200 mg | 4000 mg |
| $C_{eoi}$ (mg/L)† | 185 (74.5) [158] | 321 (106) [553] | 1049 (317) [388] | 192 (78.9) [171] | 321 (112) [580] | 1049 (308) [400] |
| $C_{28}$ (mg/L)‡ | 46.4 (22.5) [127] | 73.2 (27.2) [609] | 238 (86.1) [482] | 38.3 (19.6) [127] | 60.0 (22.9) [610] | 192 (70.2) [469] |
| Estimated $t_{1/2}$ in days (90% CI)§ | 28.8 (16.5, 41.1) | | | 25.5 (17.4, 33.7) | | |

C, concentration; eoi, end of infusion; IV, intravenous; SD, standard deviation; $t_{1/2}$, half-life.

* Mean (SD) [N], where N is number of observations
† Concentration at the end of infusion (1 hour)
‡ Observed concentration 28 days after dosing, i.e., on day 29
§ Based on 2-compartment population pharmacokinetic models developed for casirivimab and imdevimab from approximately 3700 patients across different REGEN-COV clinical trials, including this study (2067). Half-life estimates represent values for the 1200 mg IV, 2400 mg IV, and 8000 mg IV doses combined.
